# The spectrum of gene intolerance to variation: Insights from a rare disease cohort

**DOI:** 10.1101/2025.01.28.25321201

**Authors:** Pilar Cacheiro, Gabriel Marengo, David U. Gorkin, Kevin A. Peterson, Yonina Loskove, Stephen A. Murray, Damian Smedley

**Affiliations:** William Harvey Research Institute, Queen Mary University of London, London, UK; Emory University, Atlanta, GA, USA; The Jackson Laboratory, Bar Harbor, ME, USA

## Abstract

Deciphering the spectrum of intolerance to loss-of-function (LoF) variation helps identify genes that are critical at various stages of development and hierarchical levels of organisation. We have previously combined cell and mouse viability screens for single-gene knockouts, to summarise this spectrum into discrete categories, Full Spectrum of Intolerance to Loss-of-function (FUSIL), from genes essential for cell proliferation to those where LoF has no phenotypic impact. Here, we expand on this analysis to uncover distinct patterns in gene expression across developmental stages in both mouse and human within these categories, as well as gene sequence and evolutionary features, protein functional classes, and disease associations. Further, by analysing data from diagnosed patients in a rare disease cohort, we gain insights into the relationships between variant *de novo* status, molecular consequence, mode of inheritance, and type of disorder in the FUSIL categories. These associations facilitate the identification of predicted pathogenic variants in genes not currently linked to Mendelian conditions.

## Introduction

Understanding gene function and tolerance to loss-of-function (LoF) variation are two closely related objectives and often one informs on the other. Several factors complicate the assessment of these two features. Biological context, e.g. the different tissues, cell states or developmental stages in which a gene operates can impact its function^1^. Gene regulatory networks, the existence of functional redundancy or other compensatory mechanisms play a role when evaluating the phenotypic impact of a gene’s LoF, with incomplete penetrance and pleiotropy adding to the difficulty of this assessment ^2^.

Major consortia, such as the Molecular Phenotypes of Null Alleles in Cells (MorPhiC) ^3^ and the International Mouse Phenotyping Consortium (IMPC) ^4^, aim to characterise molecular phenotypes of null alleles in cell lines and in mice across life stages, including embryonic, early adult, and late adult phases. Large-scale whole-genome sequencing cohorts allow the computation of constraint metrics by comparing observed versus expected variation in the population ^5,6^. To fully understand the spectrum of intolerance to LoF variation, information from different biological levels of organisation and organisms is likely needed. We can determine whether a gene is essential for proliferation consistently across different cell lineages ^7^, whether this property can be context-dependent, or even whether it is pluripotency, rather than fitness, that is being impacted in CRISPR-Cas9 LoF screens ^8^. Similarly, in the mouse we can identify genes that are critical at different stages of embryonic development ^9^. In humans, however, cataloguing genes whose LoF leads to prenatal or neonatal death is more complex, with Mendelian forms of this extreme phenotype thought to be underrepresented in current gene-disease association resources. Several efforts are being made to address this challenge ^10,11^.

Identifying patterns of gene expression across different organisms together with genomic sequence and related evolutionary properties, can provide insights into this spectrum ^12,13^, and potentially reveal links to disease-related features.

We have previously defined the <u>Fu</u>ll <u>S</u>pectrum of Intolerance to <u>L</u>oss-of-function (FUSIL) by combining cell and mouse essentiality data and have successfully leveraged this to develop gene prioritisation approaches for developmental disorders ^14^. Assessing the functional impact of uncharacterised variants, often in genes with no previous disease associations (genes of uncertain significance, GUS) is crucial for advancing diagnostics and understanding of single-gene disorders, including those with prenatal manifestations such as foetal death. Large programs focused on the diagnosis of patients affected by rare disorders, such as the 100,000 Genomes Project (100KGP) ^15^ can be explored to identify predicted pathogenic variants in candidate genes highlighted through different prioritisation strategies.

In the present work, we update our previous FUSIL bins to incorporate mouse essentiality information from the IMPC for up to 7,000 genes. We describe how the analysis of gene expression patterns across developmental stages in different organisms uncovers associations between FUSIL bins and tissues where these genes are overexpressed or show tissue specificity. We also explore how these associations correlate with the enrichment in Mendelian genes linked to specific disease categories. Additionally, we investigate differences in sequence, evolutionary, and functional properties across FUSIL categories, as well as their relationships with patterns of human genetic variation. Lastly, we investigate both diagnosed and undiagnosed probands in the 100KGP resource to: 1) highlight patterns of pathogenic variation across FUSIL bins, and 2) identify candidate genes for different developmental disorders.

## Results

### Assessment of current FUSIL categories and follow-up on previously identified developmental lethal candidates

As new lines were systematically phenotyped through the IMPC pipeline, we now have FUSIL data for 6,905 genes. These genes have associated viability data, a minimum number of phenotypes being assessed through the broad-based phenotyping pipeline, and a one-to-one human orthologue with corresponding cell essentiality data. Our five FUSIL categories, representing different levels of intolerance to LoF variation can be summarised as follows: Cellular lethal (CL), genes essential for cell proliferation and embryonic lethal in mice; Developmental lethal (DL), genes non-essential in cell lines but embryonic lethal in mice; Subviable (SV), genes that exhibit incomplete penetrance of lethal phenotype in mouse; Viable with Phenotype (VP), homozygous viable with phenotypic abnormalities; Viable with No Phenotype (VnP), homozygous viable with no significant phenotypic abnormalities detected (normal phenotype) (**Figure 1**).

**Figure 1.**
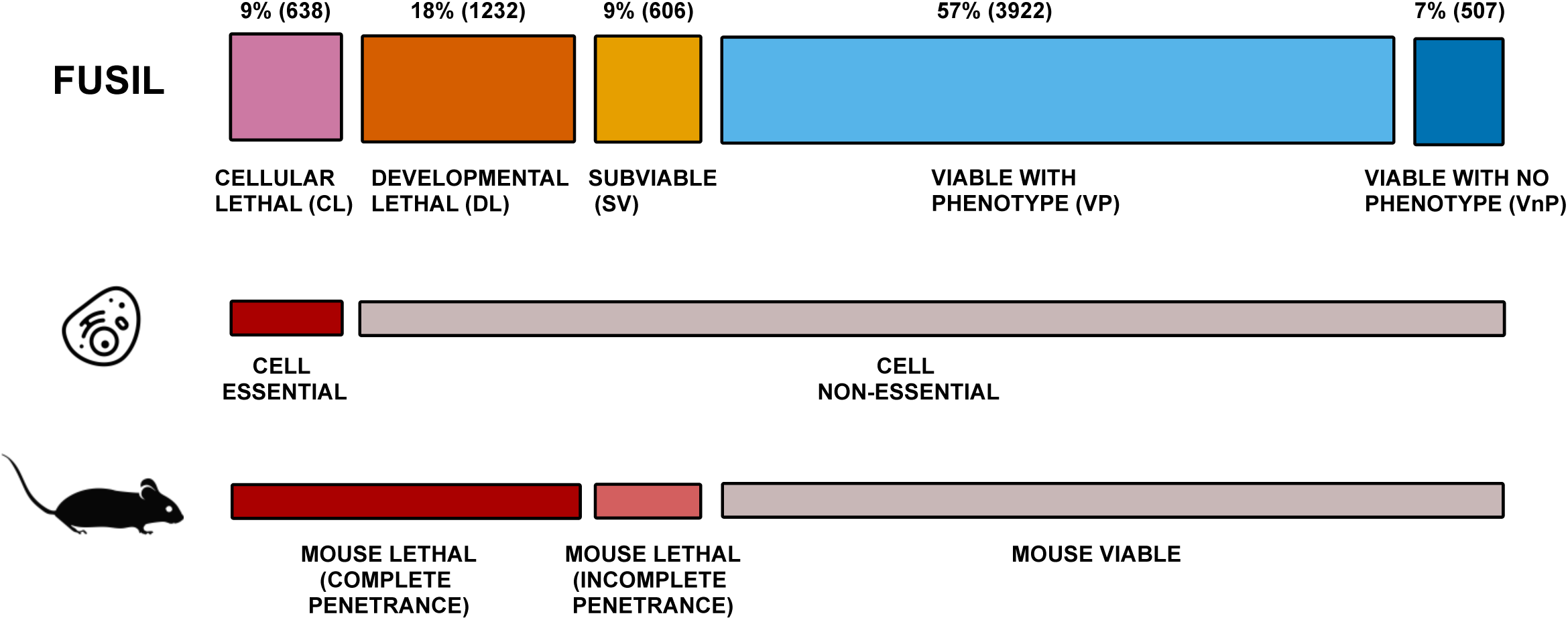
Description of FUSIL bins. Description of the FUSIL categories and the gene distribution across them. Each FUSIL bin, along with the corresponding number of genes and percentages relative to the total number of genes with FUSIL information is provided. The correspondence with categories of essentiality in cell proliferation assays and mouse viability screens is also indicated.

Our previous work focused on the enrichment of genes linked to developmental disorders within the DL bin ^14^. Based on those findings, we developed a gene prioritisation strategy that integrated constraint metrics, resulting in the identification of 163 candidate genes to be associated with developmental phenotypes in humans. Five years later, 29 of these genes have now a single-gene disorder association, and are considered diagnostic grade (green) genes by PanelApp Genomics England and/or PanelApp Australia ^16,17^. This means there is evidence from ≥ 3 unrelated families or from 2-3 unrelated families when there is significant supporting functional data. A further 7 genes have either a provisional association in OMIM ^18^ or are classified as amber in PanelApp, indicating borderline evidence for the gene-disease association. However, despite this impressive validation of the DL bin, the CL category had the highest proportion of newly discovered associations across FUSIL bins, (19% of CL that were not previously associated with disease are now green genes, compared to 18% of DL, 10% of SV, 6% VP and 4% VnP). Thus, while the DL class includes the highest overall proportion of disease-associated genes, the relatively high number of new discoveries among CL genes merits additional investigation. Consistent with previous observations, the proportion of AD-only genes is higher among the DL set: 40% vs 28%, 27%, 24% and 25% for the CL, SV, VP and VnP respectively. The neurology and neurodevelopmental disorders category (ranging from 39% to 46% across FUSIL categories) consistently had the highest number of new genes reported, followed by dysmorphic and congenital abnormality syndromes (ranging from 14% to 16%).

### Comparison with prior evaluation of FUSIL categories

As new data and improved analysis pipelines emerge, it is critical to compare our current assessment of FUSIL bins with those based on earlier human cell and mouse data releases from the same sources.

Overall, with data for approximately 3,000 new genes, we observed a slight decrease in the proportion of genes in the lethal categories, while the VP bin has increased from 49% to 57%, with the distribution of proportions across the five FUSIL bins being significantly different compared to our previous report (Chi-square p=1.28e-11). A comparison of the genes included in our previous study with the new genes added in the present work based on their knownness score ^19^, showed that the genes that entered the IMPC pipeline since 2020 are less well-characterised having a lower mean (mean=5.7, sd=7.2) compared to genes produced before 2020 (mean=7.3, sd=8.9; Wilcoxon test p <2.2e-16), which could partially explain this shift in FUSIL bins ^20^. One hypothesis is that well-known genes have stronger evidence supporting their functional or clinical relevance, making them more likely to be associated with diseases or to be lethal in mice.

A gene-based analysis to identify discrepancies in the FUSIL categories revealed that, for genes with data in both versions, 2.6% show CL-DL related fluctuations, with labels switching in both directions, 7.6% exhibit changes between VP-VnP, and 0.3% show shifts between SV and the adjacent DL and VP bins (**Supplementary Figure 1a**). A closer examination of DepMap gene effect scores for the genes that shifted between the two lethal bins showed that, as expected, their values are within the boundaries of the selected threshold (-0.5) (**Supplementary Figure 1b-c**). Data on cell proliferation for a higher number of cell lines is available, and the chosen threshold, averaged across cell lineages, may not fully capture the complexity of establishing a core set of essential genes as discussed previously ^21^. As new phenotypic assays are conducted on mouse KO lines, some genes previously categorised as VnP may now exhibit an abnormal phenotype. For lines that were previously assessed as VP but no longer show a phenotype, this change can be attributed to updates in the IMPC statistical pipeline used to assess differences between WT and KO lines, which were implemented in 2021 ^22^, as well as the inclusion of only IMPC-generated data, excluding any legacy data available in the repository. (**Supplementary Figure 1d)**. Alternative binning methods are possible, including the use of different sources of evidence, e.g. E6 self renewal scores in pluripotent stem cells ^8^, or evidence on mouse viability reported in the literature and captured by the Mouse Genome Informatics resource ^23^.

### FUSIL category genes show distinct patterns of gene expression that correlate with disease associations

To determine whether genes in different FUSIL categories have distinct patterns of expression, we first analysed RNA-seq data from the Genotype-Tissue Expression (GTEx) consortium encompassing 68 non-diseased human tissues and cell types. We calculated a gene-level Tau statistic ^24,25^ to quantify the tissue-specificity of each gene, which ranges from 0 for genes with ubiquitously consistent expression to 1 for highly tissue-specific genes. We observed a distinctive pattern of tissue specificity across the five FUSIL categories, with a clear trend from CL genes being the most ubiquitously expressed to VP and VnP showing higher tissue specificity, as indicated by their tau value distribution (**Figure 2a**). Notably, the expression level of genes within each tissue (as measured by Transcript per Million, TPM) also varies by FUSIL category, such that FUSIL categories that are more intolerant to LoF tend to have higher expression levels in a given tissue, in line with our previous report ^14^ (**Figure 2b**).

**Figure 2.**
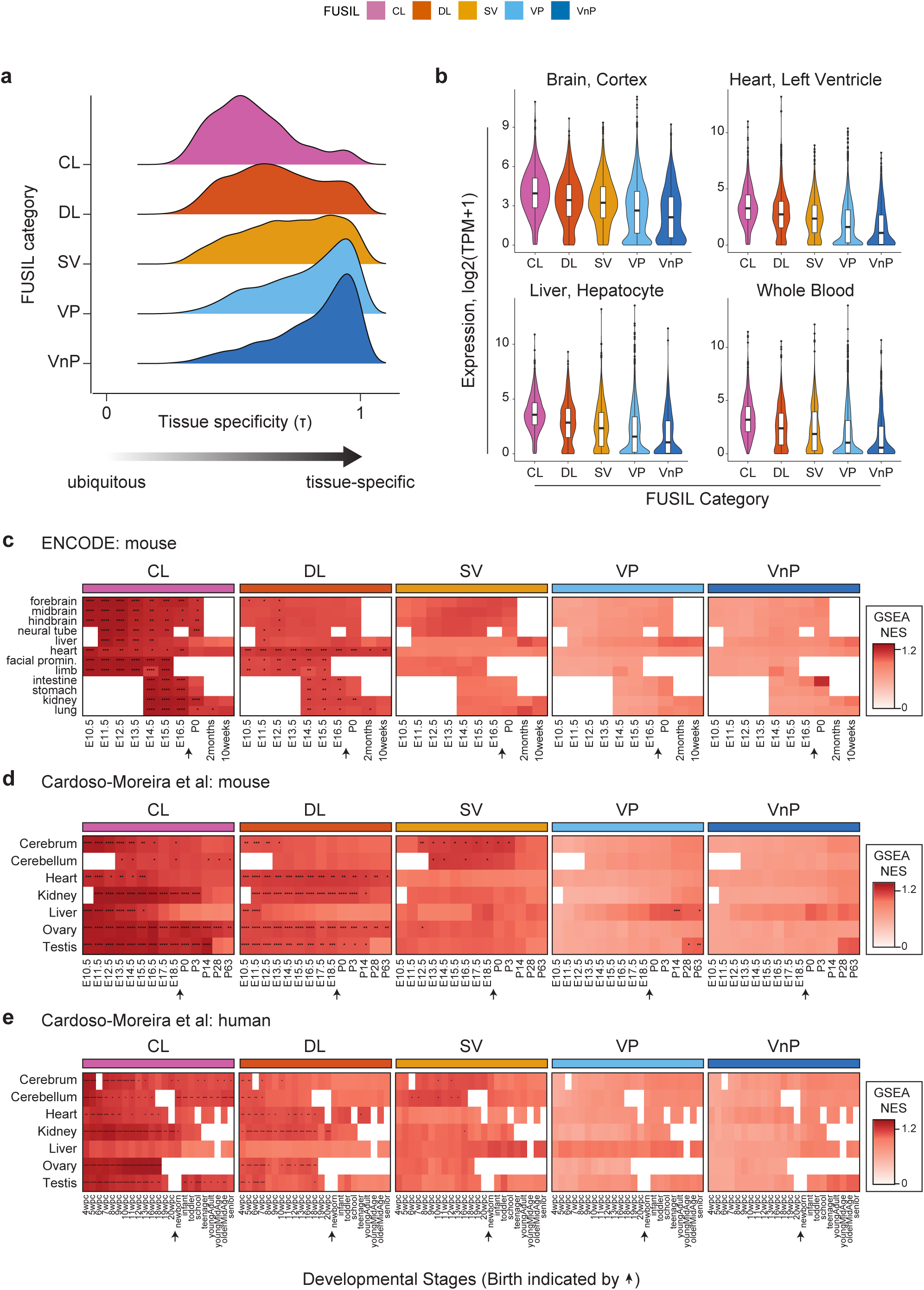
Gene expression patterns across developmental stages. (**a) Distributions of tissue specificity values (Tau) for genes in different FUSIL categories** Tau was calculated across n=68 human tissues using GTEx expression data ^63^ **(b)** Expression levels of FUSIL genes are shown in four representative tissues. TPM=Transcripts Per Million (median across tissue donors in GTEx) **(c-e) Gene Set Enrichment Analysis (GSEA)** Results of GSEA using expression data from mouse as measured by ENCODE ^27^ **(c),** mouse as measured by Cardoso-Moreira et al ^26^ **(d),** and human as measured by Cardoso-Moreira et al ^26^ **(e)**. NES=Normalized Enrichment Score. For each heatmap, the rows are tissues and the columns are developmental stages plotted in order from left to right with the time of birth indicated by an arrow head. Significant enrichments are annotated with asterisks, as follows: ****=BH adjusted p<0.0001, ***=BH adjusted p<0.001, **=BH adjusted p<0.01, and *=BH adjusted p<0.05. CL cellular lethal; DL developmental lethal; SV subviable; VP viable with phenotypic abnormalities; VnP viable with no phenotypic abnormalities.

Given that the origins of many single-gene disorders occur in early development, we next sought to examine the expression of FUSIL genes across developmental stages, including prenatally. To this end, we analysed RNA-seq data from Cardoso-Moreira et al. ^26^, which profiled a diverse panel of tissues across multiple developmental stages in both human and mouse. We also analysed a dataset from the Encyclopedia of DNA Elements (ENCODE) project that profiled multiple tissues and developmental stages in the mouse ^27,28^. Analysis of tissue specificity in these data series showed the same trend as described above for the GTEx data series, whereby FUSIL categories that are more intolerant to LoF tend to contain more ubiquitously expressed genes (**Figure 2a**). To further investigate the expression of FUSIL genes across developmental tissues and stages, we next performed Gene Set Enrichment Analysis (GSEA) ^29^ wherein genes with FUSIL annotations were ranked by their expression levels in a given tissue/stage, and then the ranks of genes in each FUSIL bin were tested for enrichment toward either the low or high of the rank distribution. A positive enrichment in this analysis means that genes in a given FUSIL bin tend to be more highly expressed in the tested tissue/stage than gene sets with randomised FUSIL annotations, and a Pvalue is calculated by permutation. This analysis revealed several interesting patterns that are consistent across all three data series examined (Cardoso-Moreira et al. human and mouse, and ENCODE mouse) (**Figure 2c-e**): First, CL genes are most strongly enriched early in development, and this enrichment is observed broadly across multiple tissues. Second, DL genes are also strongly enriched in early development, with particularly strong enrichment noted in the developing heart and kidney. Third, SV genes are most strongly enriched in the developing brain. We note that VP and VnP lack strong tissue-specific enrichments early in development, with enrichments getting marginally stronger later in development. Taken together, these results show that FUSIL genes differ at the level of expression in terms of their tissue-specificity, developmental timing, and typical levels of expression (**Supplementary Figure 2**).

We next sought to determine whether the expression patterns of FUSIL genes could provide insights into potential disease associations. Consistent with previous reports, we found that the lethal FUSIL categories (CL, DL, and SV) are associated with single-gene (Mendelian) disorders, ^14^ (**Supplementary Figure 3a**) with more specific overrepresentation of intellectual disability and neurodevelopmental disorders. Correspondingly, we observed that the lethal categories were enriched for genes with the highest relative gene expression levels in the brain, based on mouse data from Cardoso-Moreira et al. (see Methods) (**Figure 3a-b**). Developmental disorders not associated with brain and cognition were also enriched among the lethal FUSIL categories, with proportions increasing gradually from the most essential category, 33%, 40% and 43% respectively for CL, DL and SV based on the total set of developmental disease genes from DDG2P ^30^. Interestingly, genes linked to cardiomyopathies and congenital heart disease from PanelApp^14,16^ were predominantly enriched in the DL bin, with these genes showing the highest expression levels in the heart. Higher expression in the cerebellum among VP genes is supported by a higher proportion of genes associated with neurodegenerative disorders in PanelApp, although this is also observed in the CL bin (**Figure 3a-b**). The analysis of the equivalent gene expression metric (normalised TPM values, max sample (tissue+stage) for human data revealed a similar trend, although the results were not statistically significant (**Supplementary Figure 3b**).

**Figure 3.**
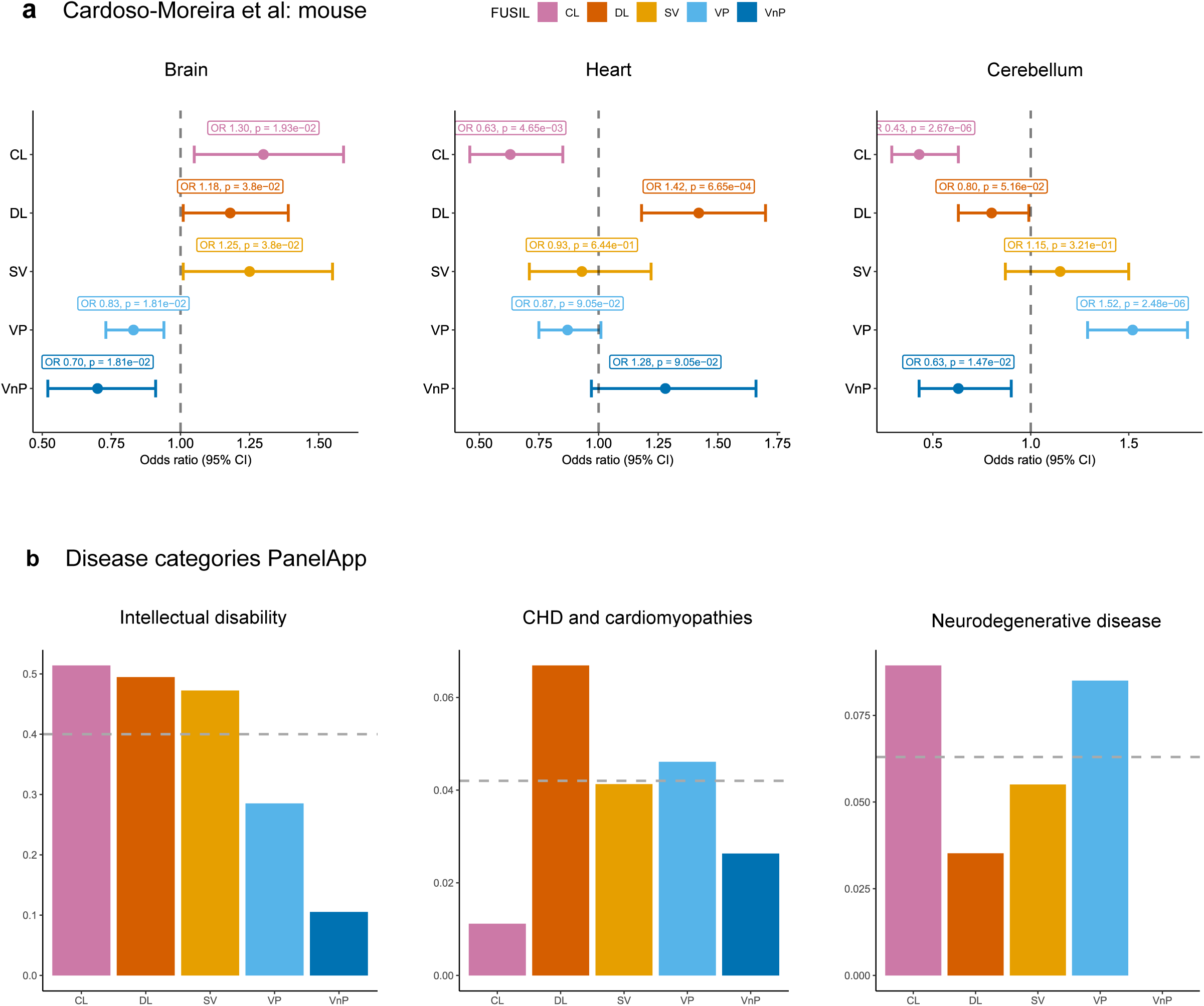
Gene expression patterns and disease associations. **(a) Enrichment analysis of tissues with the highest level of gene expression in Brain, Heart and Cerebellum** Gene expression values across mouse developmental stages were used to compute the tissue with highest levels of gene expression for each gene. Mouse data, normalised TPM values (max sample (tissue+stage)) from Cardoso-Moreira et al. ^26^ (see methods section for computation of OR). The grey dashed line represents OR=1 **(b) Distribution of genes associated with different disease categories across FUSIL bins** Bar plots represent the proportion of genes (out of all disease associated genes labelled as green - diagnostic level of evidence) included in the Intellectual Disability, Congenital Heart Disease and cardiomyopathies and Neurodegenerative disease panels in PanelApp respectively ^16,17^. The grey dashed line represents the overall proportion of genes in each panel(s) for all FUSIL bins relative to the total number of green genes in PanelApp. OR odds ratio; CHD congenital heart disease; CL cellular lethal; DL developmental lethal; SV subviable; VP viable with phenotypic abnormalities; VnP viable with no phenotypic abnormalities.

### Sequence metrics and protein classes across FUSIL bins

Based on the results from various gene expression metrics and disease associations, two distinct trends were observed across FUSIL bins: a continuous trend towards lower expression levels and greater tissue specificity, from most to least essential, and a peak in the DL and SV bins for disease-related features. These trends are consistent with the results for constraint metrics and other gene features reported in our previous study ^14^.

Here, we extended the analyses for two metrics that did not follow a trend of increasing or decreasing from most to least essential: probability of mutation and different gene length metrics. The peak in the DL and SV bins we had previously observed for transcript lengths remains consistent looking across gene, transcript, CDS and protein length, with significant differences found across non-adjacent bins (**Figure 4a**, **Supplementary Figure 4a, Supplementary Table 1**). Interestingly, when genes in each bin are categorised based on their association with Mendelian disease, only for the CL, VP and VnP bins are the transcripts of disease-associated genes longer than those of non-disease genes (**Figure 4b**).

**Figure 4.**
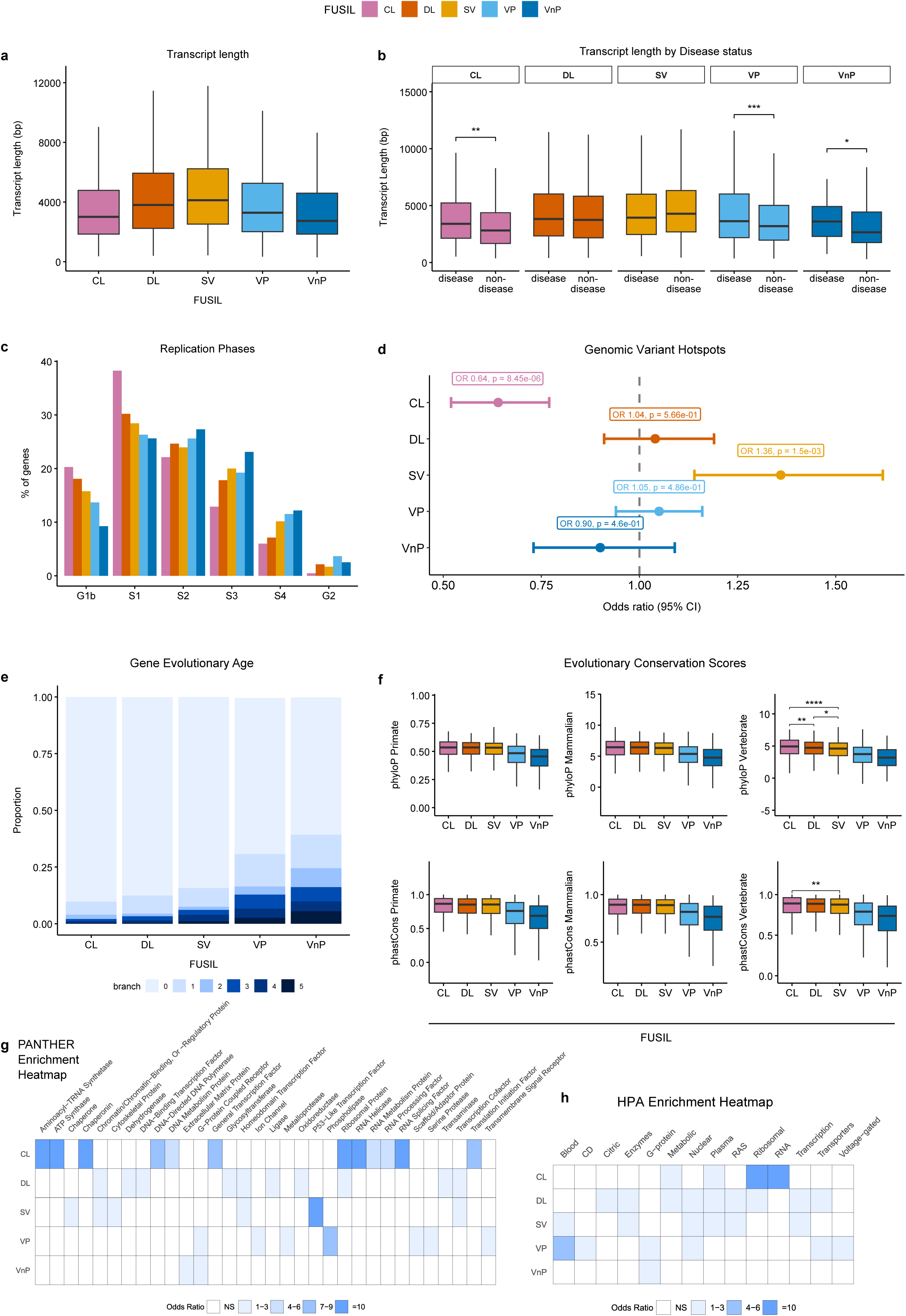
Sequence based metrics. **(a) Transcript length** Box plots show the distribution of transcript lengths (for the longest transcript for each gene) across all associated FUSIL genes (Ensembl Genes 113, hsapiens_gene_ensembl dataset) **(b) Transcript length characterised by gene disease association** Boxplots show a comparison of the distribution of transcript lengths between disease-associated and non-disease genes (determined by PanelApp Australia ^16^ and OMIM ^18^ annotations) **(c) Association with cell cycle phases** Bar plots represent the distribution of genes overlapping (≥80% overlap with cycle phase associated region) regions associated to six cell cycle phases; G1b, S1, S2, S3, S4, G2 as determined by ^31^ **(d) Enrichment of genomic hotspots** Genes overlapping (≥80% overlap with hotspot) genomic variant hotspots from Long and Xue. ^31^. The grey dashed line represents OR=1 **(e) Sequence conservation metrics** Box plots show the distribution of phyloP (top) and phastCons (bottom) scores, based on genome alignments from primates (left panel), mammals (middle panel) and vertebrates (right panel). Scores were downloaded from dbNSFP v4.7a ^35,36^. Gene level scores were computed by averaging variant level annotations. The higher the score, the more conserved the site **(f) Gene age scores** Distribution of genes across branches based on gene age prediction obtained from GenTree ^34^. The branches in the age dating system are as follows: 0 Euteleostomi; 1 Tetrapoda; 2 Aminota; 3 Mammalia; 4 Theria; 5 Eutheria **(g-h) Protein class enrichment** Heat Maps show the significant enrichment of genes associated with different protein classes derived from PANTHER ^37^ **(g)** and the Human Protein Atlas (HPA) ^38^ **(h)** in blue. White cells represent non significantly enriched protein classes (see Methods section for computation of OR, no multiple testing correction was applied across protein classes). The grey dashed line represents OR=1. For figures **(a)**, **(b)** and **(e)** outliers are not shown in the boxplots. Significant enrichments are annotated with asterisks, as follows: ****=BH adjusted p<0.0001, ***=BH adjusted p<0.001, **=BH adjusted p<0.01, and *=BH adjusted p<0.05. NS non-significant; OR odds ratio; CL cellular lethal; DL developmental lethal; SV subviable; VP viable with phenotypic abnormalities; VnP viable with no phenotypic abnormalities. The significance for pairwise comparisons across features is shown in **Supplementary Tables 1-2**.

We next explored the distribution of genes overlapping genomic regions associated with six different replication phases, corresponding to earlier and later phases of replication (G1b, S1, S2, S3, S4, G2) from Long & Xue ^31^. We observed a trend where genes in the more essential bins are increasingly skewed toward earlier replication phases, with CL being the most skewed to the earlier phases. In contrast, less essential bins gradually shift toward intermediate replication phases, with the distribution progressing from DL to SV to VP and finally to VnP (**Figure 4c**). Later replication phases are more prone to errors in replication ^32,33^, consistent with mutations accumulating in genes more tolerant to LoF. Using genomic variant (GV) hotspots from the same study, we found only SV to be enriched for GV hotspots (OR 1.36, p=2.4e-03, **Figure 4d**).

In order to better understand the relationship between mutation rates and evolutionary conservation within FUSIL bins, we then examined different metrics that evaluate gene age and conservation across different species. The analysis of the gene evolutionary age as inferred in GenTree ^34^ showed the highest proportion of older genes (branch 0, Euteleostomi) in the CL bin, decreasing gradually up to the VnP category, which in return has the higher proportion of genes in branch 5, Eutheria (**Figure 4e**). Nine different evolutionary sequence conservation scores obtained from dbNSFP ^35^ ^36^ were compared across bins (see Methods). We found that the most essential bin, CL, was the most conserved, with more tolerant to variation FUSIL genes being less conserved across all metrics. PhyloP and phastCons each have three separate metrics based on alignments of primates, mammals and vertebrate genomes respectively. Interestingly, when focusing on the lethal bins (CL, DL, SV) for these metrics, the only significant differences were found between CLvDL, CLvSV, DLvSV (phyloP vertebrate) and CLvSV (phastCons vertebrate) (**Figure 4f**, **Supplementary Table 2**). This suggests that the function of CL genes is more strongly conserved across broader evolutionary scales, while differences between lethal bins diminish at closer evolutionary distances. The distribution of different constraint metrics used to assess intolerance to LoF and missense variation, some of which have been explored previously, is shown in **Supplementary Figure 4b-c** and the significance for all pairwise comparisons across these metrics is shown in **Supplementary Table 3.**

Functional enrichment analysis of PANTHER ^37^ and The Human Protein Atlas (HPA)^38^ classes revealed differences in biological roles across FUSIL bins. Panther-enriched categories for CL proteins predominantly included essential components of transcription, translation, RNA/DNA metabolism, and protein folding such as ATP & aminoacyl-tRNA synthetases, RNA splicing factors, and chaperonins. DL proteins showed a bias towards transcriptional regulation, enzymes and metabolic processes, including DNA-binding transcription factors, glycosyltransferases, oxidoreductases and ligases. SV proteins were also skewed towards transcriptional regulation and protein folding with enrichment for P53-like transcription factors, chaperones, chromatin-binding and regulatory proteins. VP and VnP protein classes were found to be enriched for signaling pathways, including G-protein coupled receptors, ion channels, and transmembrane signaling molecules. HPA-enrichment confirmed overlapping functions, emphasising transcriptional and translational related roles in the CL bin; metabolic, ribosomal and transcription factors in the DL category; transcription factors in SV; and signalling pathway related proteins in the VP and VnP bins (**Figure 4g-h**).

### Patterns of human variation across FUSIL using diagnostic variants in the 100KGP cohort

#### Most of the diagnoses are found in DL genes

To expand beyond gene-disease associations based on OMIM and PanelApp, and to investigate pathogenic variation across FUSIL bins within a rare disease cohort, we queried the diagnostic variants in the 100KGP rare disease programme. We identified a total of 2,080 high confidence diagnostic variants in 569 unique genes with FUSIL information. Consistent with the known enrichment of Mendelian disease genes in the DL bin, 34% (n=194) of these genes fall into this category, despite representing only 18% of all genes with FUSIL data (OR 2.64, p=8.9e-22, **Figure 5a**).

**Figure 5.**
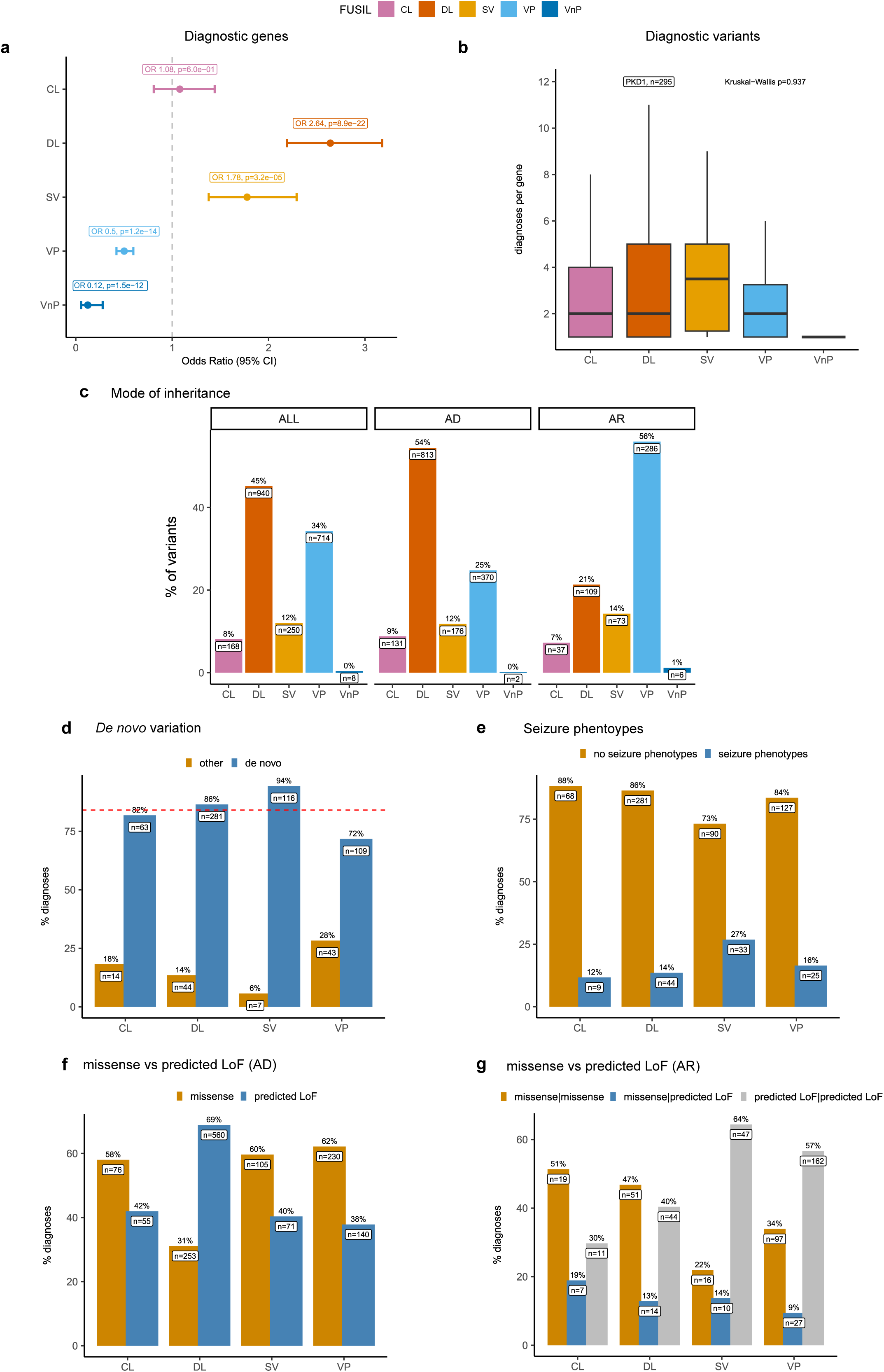
Diagnostic cases in the 100KGP. **(a) Diagnostic genes** For each FUSIL bin the number of genes with diagnostic variants in the rare disease cohort was computed to estimate Odds Ratios and 95% CI for each category. The grey dashed line represents OR=1. Adjusted BH Pvalues from the Fisher test are shown. **(b) Distribution of the number of diagnoses (variants) per gene** Box plots represent the distribution of the number of variants per gene for each FUSIL bin **(c) Distribution of diagnostic variants** Bar plots represent the number and percentage of variants in genes with FUSIL information, also categorised by mode of inheritance **(d) *De novo* diagnostic variants** Bar plots show the percentage of de novo diagnosis across FUSIL bins. Only solved cases where the family structure ≥3 (trios or larger) were considered for this analysis. The grey dashed line represents the percentage for all variants in genes with FUSIL information **(e) Seizure phenotypes** Bar plots show the distribution of *de novo* diagnosis categorised by the presence or absence of seizure phenotypes **(f) Missense versus predicted LoF variants** Bar plots show the percentage of missense and predicted LoF variants across FUSIL bins for diagnoses with an associated AD mode of inheritance **(g) Missense versus predicted LoF variants** Bar plots show the percentage of homozygous and compound heterozygous missense and predicted LoF variants across FUSIL bins for diagnosis with an associated AR mode of inheritance. For figures **(d)**, **(e)**, **(f)** and **(g)** data for VnP is not shown given the low number of *de novo* variant diagnoses in that bin (n=2). OR odds ratio; AD Autosomal Dominant; AR Autosomal Recessive; LoF loss-of-function; CI confidence intervals. CL cellular lethal; DL developmental lethal; SV subviable; VP viable with phenotypic abnormalities; VnP viable with no phenotypic abnormalities.

The analysis of the number of diagnoses per gene showed that 36% (n=295) of the diagnostic variants in the DL bin belong to one single gene, *PKD1*, associated with Autosomal Dominant (AD) polycystic kidney disease. The distribution of the number of diagnoses per gene did not show significant differences between FUSIL bins (**Figure 5b**). Exploring individual variants and their allelic requirement revealed that 45% (n=940) of all diagnostic and 54% (n=813) of all monoallelic, AD disease-associated variants when stratified by mode of inheritance are found in DL genes. Even when *PKD1* diagnostic variants are excluded from the analysis, the DL bin still contains a higher percentage of AD disease-associated variants, with 43% (n=518). In contrast, the VP bin comprises the majority of diagnostic variants associated with an Autosomal Recessive (AR) mode of inheritance, representing 56% (n=286) (**Figure 5c**).

The primary recruited disease category across FUSIL bins is intellectual disability, except in the DL category, where cystic kidney disease is the leading category. However, if diagnostic variants in *PKD1* are excluded, intellectual disability remains the dominant disease category in the DL bin as well. In both the DL and VP bins, diagnoses are more evenly distributed across disease categories, which together account for the majority of diagnoses. When broken down by inheritance mode, rod-cone dystrophy (20%, n=58) and inherited macular dystrophy (18%, n=51) are more prevalent than intellectual disability (5%, n=15) in the VP bin (**Supplementary Figure 5a**).

#### Enrichment for *de novo* mutations in SV genes

We previously reported a higher probability of mutation among the SV genes ^14^ using estimates from Samocha et al. ^39^. These results are now confirmed using different sources of evidence for mutation rates as reported in the previous section.

For the diagnostic variants where *de novo* status could be assessed, i.e. when family trios were sequenced (family structure ≥3), and with associated AD mode of inheritance, we investigated the occurrence of *de novo* pathogenic variants across FUSIL bins. Consistent with the higher mutation rates, the SV category showed a significant enrichment (SV vs non-SV OR 3.6; uncorrected p=0.00033) (**Figure 5d**).

Heyne et al. reported genes with a burden of *de novo* variants in patients with NDD with epilepsy compared to those without epilepsy ^40^. Among the 679 diagnostic variants where *de novo* status could potentially be assessed due to availability of family members, 111 (16%) were associated with seizure phenotypes. The SV bin was found to be significantly enriched for this phenotype (SV vs non-SV OR 2.24; uncorrected p=0.00108). Consistently, *de novo* variants associated with seizure phenotypes were enriched in the SV bin (SV vs non-SV: OR 2.04; uncorrected p=0.0047). No seizure phenotypes were observed among the non-*de novo* variants (n=7) in this category (**Figure 5e**).

#### Missense vs predicted LoF across FUSIL categories

The analysis of the variants’ functional class revealed that, for monoallelic variants, i.e. associated with AD inheritance, the DL bin is depleted for missense variants. This is the only category with a lower percentage of missense variants compared to predicted LoF variants, making it distinct for this feature (CL: 58%; DL: 31%; SV: 60%, VP: 62%, VnP: only 2 variants). This remains true even when variants in *PKD1* are excluded (41%). For homozygous and compound heterozygous variants, the percentage of predicted LoF as opposed to missense variants are as follows: (CL: 30% ; DL: 40%; SV: 64%, VP: 57%, VnP: only 5 variants). The depletion of homozygous pLoF variants in the two lethal bins is consistent with null alleles being lethal in mice, however, discrepancies persist between mouse and humans as postnatal (viable) phenotypes are observed in patients. Previous reports have highlighted these differences in lethality between null alleles in mice and humans, offering potential explanations, which will be discussed further later ^9,11^ (**Figure 5f-g**).

#### Patients phenotypic similarity within and across FUSIL categories

To explore potential associations between genes within the same FUSIL bin and similar phenotypes, we performed pairwise comparisons of phenotypic similarities for all individuals with a diagnosis in genes with FUSIL information. Phenotypic similarity algorithms allow us to computationally assess the similarity of clinical manifestations between patients, using pairwise similarity scores derived from their associated phenotypes, encoded as HPO terms ^41^. This helps determine whether patients with pathogenic variants in genes within the same FUSIL bin exhibit higher phenotypic similarity compared to those with pathogenic variants in genes from different FUSIL bins. After excluding pairwise scores for patients diagnosed with the same gene, distinct patterns across FUSIL bins emerged.

The expected pattern where similarity scores for patients diagnosed with genes in the same FUSIL bin for the same disease > different FUSIL bin same disease > same FUSIL bin different disease > different FUSIL bin different disease largely holds true for the VP bin and partially for the DL bin, where patients affected by the same disease and a diagnoses in a gene in the same bin showed higher pairwise phenotypic similarity scores compared to those with a pathogenic variant in a gene located in a different FUSIL bin (**Supplementary Figure 5b**). In contrast, for the CL and SV bins, the disease category is the main discriminating factor, and having a diagnosis in a gene within the same FUSIL bin does not significantly affect the phenotypic similarity between patients. This may suggest that the genes in these bins are more heterogeneous in terms of their associated phenotypic outcomes, with CL genes likely having broad, pleiotropic effects with significant context-specific modification.

#### Novel candidate gene discovery in the 100,000 Genomes Project

Building on our previous work ^14^, we applied a similar approach, leveraging both the new genes with FUSIL information that are not currently associated with disease and the consistent findings regarding enrichment in Mendelian disease genes (DL, SV, particularly in the case of AD disorders), overrepresentation of *de novo* variants (SV), and associations with relevant tissues (brain in CL,DL,SV; heart in DL). Based on this, we explored different strategies to identify potentially pathogenic variants in patients who remain undiagnosed in the 100KGP. We focused on genes in the DL and SV bins with no established Mendelian disease association, where we identified rare variants with an inheritance pattern consistent with family history that were either i) *de novo* or ii) ranked among the top 5 Exomiser variants with an associated AD mode of inheritance. In both scenarios, we focused on genes where at least two patients, recruited under the same disease category, carried that met the above criteria, and where no variants meeting the same criteria were found in patients from other disease categories.

Genes with variants matching these criteria were assessed using the ClinGen framework ^42^ (see Methods) which includes a genetic score based on the type of variation besides scores for gene function, gene expression, protein-protein interaction, and animal model evidence. Additionally, evidence of disease associations for other genes in the same gene family, the presence of variants being classified as research variants in Decipher ^30^, predicted probabilities from Mantis-ml ^43^ and multiple constraint metrics were considered. IMPC heterozygous knockout mice ^4^ were queried to determine if they had associated abnormal phenotypes mimicking those observed in patients. Evidence for the clinical validity assessment was considered moderate for *HDAC2*, *and LHX6*. For the remaining genes, the evidence was scored as limited.

In *HDAC2*, the top ranked gene by the ClinGen framework, two *de novo* frameshift and two *de novo* missense variants are ranked 1,1,2,2 respectively by Exomiser in three patients with intellectual disability and one with osteogenesis imperfecta. *HDAC2* has been recently highlighted as a potential causative gene for chromatinopathies ^44^ and featured as a candidate gene in a combined analysis of single-nucleotide and copy number variants in neurodevelopmental disorders ^45^. The Mantis score is 99.5 and it is classified as highly intolerant to heterozygous variation by LOEUF. Two different *de novo* missense variants are also reported in the Decipher Research Variants set. Several other genes from the same family (*HDAC3*, *HDAC4,* and *HDAC8*) are green (diagnostic) genes in the PanelApp Intellectual Disability (ID) panel. Interestingly, the heterozygous IMPC mouse shows no spontaneous movement and unresponsive to tactile stimuli phenotypes.

*LHX6* is the second-highest ranked gene based on the ClinGen framework, the evidence was more limited, with some functional data primarily coming from cell line studies ^46,47^. Two patients with epilepsy have one start loss and one splice variant ranked 2,1 by Exomiser. The IMPC heterozygous knockout shows decreased locomotor activity. The gene gets a mantis score of 96.7 and is also considered intolerant to heterozygous LoF variation according to different constraint metrics. Within the gene family, we find *LHX2* as a diagnostic-grade gene in the ID panel, and *LHX3* and *LHX4* as diagnostic-grade genes in the growth failure panel.

*LEO1,* even though only scoring as limited evidence, has been recently identified through a burden re-analysis of neurodevelopmental disorder cohorts by the UDN program, where they collated additional evidence on other *de nov*o and inherited variants associated to ID/ASD phenotypes ^48^. Two undiagnosed patients in the 100KGP cohort carry *de novo* missense variants ranked 3 and 11 by Exomiser, although *de novo* variants are also present in one patient with hereditary ataxia and one with intracerebral calcification. This gene is also highly intolerant to variation according to multiple metrics, gets a Mantis-ml score of 93.5 for ASD and is currently labelled as red in the PanelApp Autism panel. Another gene with limited evidence^49^ is *DAPK3,* with two *de novo* missense variants ranked 2,3 by Exomiser, identified in patients with intellectual disability. The heterozygous IMPC mouse shows hyperactivity, however this gene is considered tolerant to heterozygous LoF based on its LOEUF score.

Within the 100KGP cohort, cardiomyopathies and congenital heart disease constitute disease categories with a lower number of recruited patients, making it more challenging to find variants meeting the criteria described above, specifically, candidate genes in the DL bin with at least two undiagnosed patients carrying highly predicted pathogenic variants in the same gene. A missense variant with an Exomiser variant score of 0.95 (ranked 2) in *GJC1* was found in a patient with dilated cardiomyopathy. The IMPC homozygous model shows abnormal pericardium morphology in the early embryonic developmental stages (LOEUF=0.348). Previous studies found *GJC1* to be associated with atrial fibrillation and congenital heart disease ^50,51^. The candidate variants and supporting evidence for all these genes is summarised in **Supplementary Table 4**.

## Discussion

The investigation of the full spectrum of intolerance to variation, by integrating data across species and biological levels of organisation, continues to uncover new associations with disease-related features and guide the prioritisation of candidate genes and variants. Lethal genes in the mouse remain a valuable source for identifying novel disease candidate genes. However, as we obtain viability data for less-studied, less-essential genes ^19,20^, the proportion of lethal genes may ultimately be lower than previous and current reports suggest. When lethal genes in the mouse are categorised by their essentiality status in cell lines, DL genes, those non-essential in cell lines, retain the highest enrichment for disease-associated genes, particularly AD disorders. However, CL genes, those lethal in mice and essential in cell lines, should not be overlooked, since they have led to the highest proportion of novel discoveries since our previous publication.

These categories are not rigid, as evidenced by a small percentage of genes shifting between bins since our first study. As new data emerge, some fluctuations between adjacent bins are expected. For example, the distinction between CL and DL genes is based on a threshold applied to a continuous score. Nonetheless, this categorisation allows us to capture and simplify a continuum, and the distinction between these two types of lethal genes drives distinct trends across FUSIL categories, evident in sequence metrics, expression patterns, and protein enrichment. Discrepancies between species exist ^11^, and the analysis of the different bins from an evolutionary perspective suggests that the CL bin may contain essential genes common across multiple species, whereas genes critical for organism development in the DL bin could vary between species.

Unsurprisingly, CL genes are depleted for genomic variant hotspots and most abundant in the earlier (G1b & S1) phases of the cell cycle, which are less prone to mutations ^32,33^, highlighting their intolerance to genetic variation. CL genes are also more conserved across vertebrate genomes, although this trend weakens in mammalian or primate-specific comparisons. Similarly, Jordan et al. ^52^ found that orthologues of essential *E. coli* genes are more broadly distributed among bacterial and archaeal species than orthologues of nonessential *E. coli* genes. Supporting this, early mouse lethal genes, where the embryo dies before embryonic day 9.5, and which highly overlap with CL genes, exhibit a higher proportion of paralogues originating from the broad eukaryotic clade Opisthokonta compared to Bilateria ^9^, a more recent evolutionary clade. This widespread phylogenetic distribution and conservation of genetic sequence across species indicates ancient origins of CL genes. In fact, the CL category shows the highest proportion of genes predicted to have originated earlier in evolutionary time ^53^.

The impact of LoF on organismal viability and fitness is crucial for understanding gene molecular function. However, other factors, such as the influence of LoF on pluripotency, may be equally significant ^8^. This is particularly relevant for the DL bin, where genetic perturbations affect multicellular level processes, highlighting the necessity to explore how developmental trajectories are impacted by genomic variation. The enrichment of DL genes in proteins associated with transcriptional regulation, extracellular matrix components, and other developmental pathways underscores their role in orchestrating these trajectories, highlighting their importance in regulating complex processes such as cellular differentiation and tissue development which are crucial for organism development.

The peak in coding length observed in DL and SV bins is consistent with expression of the longest human genes being enriched in neuronal tissues ^54^ and the enrichment for NDD associated genes observed in the DL bin. However, when splitting by disease association status, it is precisely in these two bins where we do not find statistically significant differences in coding length between disease-associated and non-disease genes regarding coding length. We need to understand how disease associations and their underlying disease mechanisms relate to genes in different intolerance bins, and thus with differing impact on organismal viability. For instance, essential genes often function as hubs within protein-protein interaction networks ^12^, where disruptions can trigger cascading effects that indirectly contribute to disease.

In-depth analysis of gene expression patterns across organisms and developmental stages shows that expression levels and breadth of expression show distinct patterns across FUSIL categories. This is fairly consistent across organisms and datasets. When this is coupled with disease gene associations, patterns in tissues match enrichment for disease categories. Computational approaches are being developed that allow to project genetic associations in complex traits through patterns of gene expression, revealing modules of genes with similar expression profiles in specific tissues and cell types across similar disorders ^55^. In a similar context, characterising gene expression profiles in relevant tissues can inform potential follow-up functional studies ^56^. Thus, mapping gene expression patterns to disease associations can help prioritise candidate variants and genes based on the observed phenotypes.

The association between genes essential for organism development and genes associated with Mendelian disorders remains solid and consistent as new data on mouse viability becomes available and novel gene-disease associations are uncovered. While the DL bin continues to be the category with the highest enrichment, and consequently, a reservoir for new candidate genes, particularly monoallelic forms of developmental disorders, the findings observed for the CL bin in terms of novel gene discoveries, as well as the SV bin when investigating the FUSIL spectrum in the context of a rare disease cohort, suggest that these two categories deserve more attention. In particular, the SV category, characterised by incomplete lethal phenotypes in mice, with its distinct features, emphasises the need to understand the mechanisms behind variable penetrance. These mechanisms, along with factors like expressivity and pleiotropy, complicate the assessment of the pathogenicity of genetic variants, and consequently, the molecular diagnosis ^2^.

Understanding the relationship between longer transcripts, higher mutation rates, and disease associations is challenging, with emerging evidence indicating that certain phenotypes may be more frequently observed in diagnoses due to *de novo* variation ^40^. Tools that allow for an automated genotype-phenotype correlation analysis in cohorts of patients will facilitate the identification and/or confirmation of new and existing patterns. Given the observed association between mutation hotspots and the occurrence of pathogenic variants, resources capturing this information are crucial to facilitate the diagnosis of Mendelian disorders beyond cancer genomics ^57^.

The analysis of FUSIL categories in the context of a rare disease cohort allowed us to explore correlations at the variant level, revealing discrepancies in the phenotypic impact of homozygous LoF variation between mouse and human. We observed a higher ratio of homozygous missense variants to homozygous LoF variants in humans in genes where complete knockouts lead to embryonic lethality in mice, suggesting that homozygous LoF variants in these genes could also be lethal in humans. In contrast, this ratio is reversed in genes that are viable in mice. Homozygous LoF variants are associated to non-lethal postnatal phenotypes in humans for mouse lethal genes, and viable mouse knockouts have also been associated to early lethal phenotypes in humans ^9,11^. Here, it is important to consider potential species-specific physiological differences and potential gene function differences that may result in changes to gene regulatory network architecture. These differences can be tested by leveraging genetic diversity resources available in the mouse. All IMPC data is collected on a C57BL/6N genetic background which allows for cross-comparison across different phenotyping centers; however, there is ongoing work to gain a better understanding of how genetic diversity in mouse strains can be used to model specific human diseases.

Our previous work was highly successful in identifying novel candidate genes for neurodevelopmental disorders, by identifying *de novo* missense variants in DL candidate genes in patients with similar phenotypes ^14^. Several years later, and thanks to all the efforts in advancing diagnosis in the 100KGP ^15^, the likelihood of identifying predicted pathogenic variants in the same gene across multiple undiagnosed patients with neurodevelopmental phenotypes has decreased. However, some variants highly ranked by Exomiser are located in genes with moderate functional evidence, making them strong candidates for investigations in other cohorts.

Although the primary focus of this study has not been on the less essential VnP bin, we want to emphasise the significance of this category of genes. Despite the limitations in assessing this category, which is limited to the phenotypes captured by IMPC assays, and with all the associated methodological constraints, certain patterns are consistent and significant. The absence of molecular diagnoses in the 100KGP in genes within this category, combined with the observation that these genes are less constrained, suggests they may be dispensable or genetically redundant. Further investigation of these genes in human population cohorts where homozygous LoF variants with no clinical manifestations are found will help us understand the mechanisms underlying the absence of observable phenotypes ^58^.

Overall, our FUSIL bins are derived from single-gene experiments that assess the dependency of a cell line or organism viability on a specific gene, while genes may have multiple functions depending on the biological context. One limitation in addressing this pleiotropy is the structure of ontologies of functions, which drives the exploration of alternative strategies to map genes and their possible functions across different contexts ^1^. Despite these challenges, our FUSIL categories provide a framework for understanding the continuum of intolerance to LoF variation. Categorising genes into distinct groups based on various thresholds or criteria contributes to shedding light on the intricate variability in gene intolerance to variation, a challenge that the human genetics community continues to address. Beyond their direct application in assisting the diagnosis of single-gene disorders, these categories may also prove valuable for drug target discovery and safety assessment, for example when investigating the properties of genes where LoF leads to abnormal phenotypes, but pharmacological intervention is tolerated ^59^.

## Methods

### FUSIL categories

Mouse data on viability was obtained from the IMPC viability report (DR 21.1) ^4^ [https://ftp.ebi.ac.uk/pub/databases/impc/all-data-releases/release-21.1/results/, file: viability.csv.gz; Data accessed 24.07.24]. Further information on this procedure is available through the IMPC website, including the description of the viability primary screen procedure (https://www.mousephenotype.org/data/embryo).

Mouse gene-phenotype associations were obtained from the IMPC genotype-phenotype assertions report (DR 21.1) [https://ftp.ebi.ac.uk/pub/databases/impc/all-data-releases/release-21.1/results/; file: genotype-phenotype-assertions-IMPC.csv.gz; Data accessed 24.07.24]. No genotype-phenotype assertions from legacy data were used (e.g. 3I, MGP, EUROPHENOME). Information of procedures completeness was obtained from the procedure completeness report [https://ftp.ebi.ac.uk/pub/databases/impc/all-data-releases/release-21.1/results/; file procedureCompletenessAndPhenotypeHits.csv.gz; Data accessed 24.07.24). To further categorise the viable genes into VP and VnP, we first assessed whether a phenotype was observed in the early adult mice. For those without a phenotype hit, we evaluated whether the number of successful procedures in the homozygous or hemizygous early adult mice met a threshold. Genes with 13 or more successful procedures and no abnormal phenotypes were categorised as VnP, while lines with no abnormal phenotypes and fewer than 13 successful procedures were excluded from the final FUSIL set.

CRISPR Gene Effect scores were obtained from DepMap ^7^ (DepMap 24Q2 Public) [https://plus.figshare.com/articles/dataset/DepMap_24Q2_Public/25880521; Data accessed 24.07.24]. Gene symbols were mapped to HGNC IDs ^60^. A similar approach to that used in Cacheiro et al. ^14^ was applied to establish a cutoff for cellular essentiality, based on the average score across the 1150 cell lineages available, with a threshold of -0.5.

Mouse-human orthologue mapping files are generated weekly as part of the internal IMPC pipeline to monitor the production and phenotyping of mutant mice in the Genome Targeting Repository. These include one-to-one orthologue calls that were used to assess the FUSIL categories [https://www.gentar.org/orthology-api/api/ortholog/one_to_one/impc/write_to_tsv_file; Data accessed 24.07.24]. This analysis is limited to protein-coding genes.

### Genes associated to human single-gene disorders

OMIM [https://omim.org/] ^18^, DDG2P [http://www.ebi.ac.uk/gene2phenotype/] ^61^, PanelApp Genomics England ^16^ [https://panelapp.genomicsengland.co.uk/] and PanelApp Australia ^17^ [https://panelapp.agha.umccr.org/] were queried to verify the current status of the set of DL genes with no evidence of Mendelian phenotype association back as of 2020 [Data accessed 04.11.2024]. Data from previous FUSIL assessment was obtained from Cacheiro et al.^14^. These Mendelian genes are assigned to disease categories based on Level 2 and Level 3 panel data to identify panels linked to specific disorders [https://panelapp.genomicsengland.co.uk/; https://panelapp.agha.umccr.org/ ; Data accessed 04.11.24]. These annotations were used for the analysis shown in Figures 3a, 3b, 4b and Supplementary Figures 3a and 3b.

### Gene expression analysis

GENCODE annotations were used as a reference gene list onto which we could map expression data from different studies (v47 for human, vM36 for mouse). For each expression dataset, we used gene-level quantifications from the original study, and matched genes to the GENCODE reference by Ensembl gene ID. For GTEx data, we used median Transcripts Per Million (TPM) across individuals as provided through the GTEx portal. For the expression datasets from Cardoso-Moreira et al., expression measurements were provided as Reads Per Kilobase Million (RPKM). We averaged RPKM values across biological replicates to produce a single quantification per gene per tissue and stage. For ENCODE data, we used gene-level TPM values calculated by the ENCODE uniform processing pipeline from pooled replicates and available through the ENCODE portal.

To quantify the tissue specificity of each gene we calculated Tau values for each as previously described ^24^ and benchmarked. We performed Gene Set Enrichment Analysis (GSEA) to test for enrichment for FUSIL genes among those highly expressed in each tissue using an implementation in the FGSEA Bioconductor package ^62^. Briefly, genes were ranked by expression in a given tissue-stage, and an enriched score was calculated for each set of FUSIL genes using a running-sum statistic across the ranked list. This enriched score was then normalised relative to 1000 randomised gene sets to generate a Normalized Enrichment Score (NES) that can be compared across FUSIL gene sets. Pvalues are calculated using a Monte Carlo estimation of the frequency with which a randomized gene set of similar size would show a similarly extreme enrichment score, as implanted in FGSEA. Pvalues shown here are corrected for multiple testing of multiple FUSIL categories using the Benjamini–Hochberg procedure. The following datasets are used in the text and figures: GTEx ^63^ (Human) [https://gtexportal.org/home/downloads/adult-gtex/bulk_tissue_expression; file GTEx_Analysis_v10_RNASeQCv2.4.2_gene_median_tpm.gct.gz, Data accessed 06.01.25]; Cardoso-Moreira et al ^26,63^ (Human) [https://www.ebi.ac.uk/biostudies/arrayexpress/studies/E-MTAB-6814; file Human.RPKM.txt; Data accessed 12.01.25]; Cardoso-Moreira et al ^26,63^ (Mouse) [https://www.ebi.ac.uk/biostudies/arrayexpress/studies/E-MTAB-6798; file Mouse.RPKM.txt; Data accessed 12.01.25]; ENCODE ^27,28^ (Mouse) [https://www.encodeproject.org/ Data accessed 17.01.25]; The full list of ENCODE files used are provided in **Supplementary Table 5**.

### Sequence and functional annotations

Transcript lengths were obtained using the biomaRt R package (Ensembl Genes 113, hsapiens_gene_ensembl dataset)^64^. For each HGNC ID, the maximum transcript length was calculated across all associated gene transcripts. Gene lengths were obtained using the biomaRt R package (Ensembl Genes 113, hsapiens_gene_ensembl dataset)^64^. For each HGNC ID, the most recent Ensembl Gene ID version was selected. Coding DNA Sequence (CDS) lengths, the portion of a transcript that excludes UTRs, were obtained using the biomaRt R package (Ensembl Genes 113, hsapiens_gene_ensembl dataset)^64^. For each HGNC ID, the MANE transcript was selected, if no MANE transcript was available, the longest CDS was selected. Protein lengths were obtained from the UniProtKB REST API (Human data set) using the queryup package ^65^. The longest protein length was selected.

Genomic Variant (GV) hotspots and genomic regions associated with replication phases were obtained from Long and Xue ^31^. The HelloRanges^66^ and GenomicRanges^67^ R packages were used to map genomic regions to genes, with regions assigned to genes if 80% or more of their length overlapped a gene.

Evolutionary conservation metrics (phyloP and phastCons) from dbNSFP^35, 36^ v4.7 were averaged across all transcripts per gene using R v4.3.0 to calculate gene-level scores [Data accessed 9.09.24]. Gene evolutionary age was obtained from GenTree ^53^ [http://gentree.ioz.ac.cn/; Data accessed 10.01.25].

PANTHER^37^ protein classes were obtained using the PANTHER.db^68^ R package. Human Protein Atlas (HPA) protein annotations were obtained from the HPA webportal [Data accessed 13.11.24].

gnomAD LOEUF (loss-of-function observed/expected upper bound fraction) and Missense Z scores were obtained from the gnomAD v4.0 (https://gnomad.broadinstitute.org/) ^69^ [Data accessed 07.01.2024]. Constrained genes are defined using the cut-off LOEUF < 0.6. GISMO and GISMO-mis (missense) scores were obtained from Liao et al. ^70^. S_het_ (GeneBayes) metrics, derived from the machine learning model GeneBayes, were obtained from Zeng et al. ^71^. AlphaMissense pathogenicity scores were obtained from Cheng et al ^72^. For each HGNC ID, the MANE transcript was selected, if no MANE transcript was available, the Canonical transcript was used instead.

### Computation of Odds Ratios

To compute the enrichment of different features as shown in Figures 3a, 4b, 4g, 4h, 5a, and Supplementary Figure 3b we used only genes with FUSIL data. For each gene bin (X) and feature (Y), a 2x2 contingency table is created with the following categories: genes in bin X annotated with feature Y, genes in bin X not annotated with feature Y, genes not in bin X annotated with feature Y, and genes not in bin X not annotated with feature Y. The odds ratio (OR) is then calculated to assess the enrichment of feature Y in bin X, and Fisher’s Exact Test is used to determine if this association is statistically significant. This process is repeated for every bin and feature. ORs were computed using the epitools^73^ R package and calculated by unconditional maximum likelihood estimation (Wald). Pvalues shown are corrected for multiple testing across the FUSIL categories using the Benjamini–Hochberg procedure. Yates’s correction for continuity was applied when no viable contingency table was available.

### Analysis of diagnostic variants in the 100KGP cohort

The analysis of diagnostic variants in FUSIL genes from the 100,000 Genomes Project rare disease cohort was performed in the Genomics England Research Environment, accessing the data in the National Genomic Research Library, managed by Genomics England ^74^. A description of the rare disease cohort, including genome sequencing analysis is provided by Cipriani et al ^75^. This analysis was limited to rare disease solved cases, i.e. those patients where a diagnostic variant has been reported. Diagnostic variants mapping to genes with FUSIL information were included for subsequent analysis. Information on diagnosis status was updated on 20.11.2024 with information from Genomics England LabKey (gmc exit questionnaire and submitted diagnostic discovery). Information on recruited disease category, variant functional class and zygosity, family structure and *de novo* status were used for this analysis. Variant annotation information was obtained with Exomiser (see next section). Phenotype similarity scores were computed using the PhenoDigm, based on the associated HPO terms for each patient.^41^ Pairwise scores for patients with a diagnostic variant in the same gene were excluded.

### Novel gene discovery in the 100KGP

Exomiser (version 12.1.0 with default settings and latest databases (July 2020)) ^76^ was run on single proband and family-based variant call format (VCF) files to obtain a pool of rare protein-coding variants (< 0.1% autosomal/X-linked dominant or homozygous recessive, < 2% autosomal/X-linked compound heterozygous recessive based on gnomAD). Segregating and predicted pathogenicity filters (based on REVEL and MVP) were also applied. Likely false positive or cohort-specific variants were further removed by filtering based on how often they were observed in the Exomiser final results for the whole of the 100KGP (frequency > 2% for variants in a compound-heterozygote genotype, > 0.2% for mtDNA genome variants, > 0.1% for heterozygote/homozygote variants).

ClinGen criteria classification was applied to assess the strength of gene-disease associations (https://www.clinicalgenome.org/site/assets/files/9232/gene-disease_validity_standard_operating_procedures_version_10.pdf). A genetic evidence score was calculated from scoring and summing all candidate variants for each gene (1.5 points for LoF variants or 2 if *de novo*, whilst others scored 0.1 points or 0.5 if *de novo*) capped at a maximum of 12. Existing evidence for the gene-disease association in PubMed was assigned a score of 0, 1 or 2 for none, some tenuous or lots of evidence respectively. A score of 0, 1 or 2 was assigned for no, widespread or solely specific expression in the relevant disease tissue, based on gene expression evidence from the GTEx Project which was accessed through the web portal of the Human Protein Atlas (https://www.proteinatlas.org/) ^38^. Defaults of one point for protein-protein association evidence (high quality, direct experimental interactions scoring > 0.7 in StringDB with genes on the disease panel from PanelApp 14) and 2 points for mouse evidence (phenotypic similarity as calculated by Exomiser between the patient’s phenotypes and the mouse phenotypes where the orthologous gene was disrupted) were used. The rounded sum of genetic and experimental evidence points was used to assign the final ClinGen classification of the evidence for the association as being either limited (0.1-6 points), moderate (7-11 points) or strong (12-18 points).

## Data availability

Research on the de-identified patient data used in this publication can be carried out in the Genomics England Research Environment subject to a collaborative agreement that adheres to patient led governance. All interested readers will be able to access the data in the same manner that the authors accessed the data. For more information about accessing the data, interested readers may contact or access the relevant information on the Genomics England website: https://www.genomicsengland.co.uk/research.

## Acknowledgements

This research was made possible through access to data in the National Genomic Research Library, which is managed by Genomics England Limited (a wholly owned company of the Department of Health and Social Care). The National Genomic Research Library holds data provided by patients and collected by the NHS as part of their care and data collected as part of their participation in research. The National Genomic Research Library is funded by the National Institute for Health Research and NHS England. The Wellcome Trust, Cancer Research UK and the Medical Research Council have also funded research infrastructure.

## Contributions

P.C., G.M., D.U.G. and D.S. contributed to data analysis, data/results interpretation, writing the manuscript, reviewing and editing the manuscript. K.A.P. contributed to data generation, data/results interpretation, reviewing and editing the manuscript. Y.L contributed to data analysis, data/results interpretation, reviewing and editing the manuscript. S.A.M. contributed to data/results interpretation, reviewing and editing the manuscript. P.C., S.A.M. and D.S. are PIs of a key program who contributed to the management and execution of the work.

The authors declare no competing interests.

