## Supplementary Information for "The spectrum of gene intolerance to variation: Insights from a rare disease cohort"

Cacheiro et al.

Content

**Supplementary Figures**

Supplementary Figure 1 Comparison with previous FUSIL assessment

Supplementary Figure 2 Distributions of tissue specificity values (Tau)

Supplementary Figure 3 Gene expression patterns

Supplementary Figure 4 Sequence length and constraint metrics

Supplementary Figure 5 Diagnostic cases in the 100KGP

**Supplementary Tables**

Supplementary Table 1 Gene length

Supplementary Table 2 Sequence conservation metrics

Supplementary Table 3 Constraint metrics

Supplementary Table 4 Candidate variants identified in DL and SV genes

Supplementary Table 5 ENCODE file information

**Supplementary References**

#### Supplementary Figure legends

**Supplementary Figure 1 Comparison with previous FUSIL assessment**

**(a) Gene reclassification between FUSIL assessment** An alluvial plot shows how genes are labeled in different categories between the previous [1] and current FUSIL assessments **(b) Distribution of DepMap scores for genes that moved between the CL and DL bins** Boxplots show the distribution of mean DepMap CRISPR gene effect scores for genes that moved from the CL to the DL bin and from the DL to the CL bin**.** The grey dashed line represents the threshold (-0.5) used to categorise mouse lethal genes into the CL and DL bins. For the CERES scores, more negative values indicate stronger growth inhibition [2] **(c) Distribution of DepMap scores across FUSIL categories** Boxplots show de distribution of mean DepMap CRISPR gene effect scores across FUSIL categories. The grey dashed line represents the threshold (-0.5) used to categorise mouse lethal genes into the CL and DL bins **(d) Genes that moved between the VP and VnP bins** The number of genes that move between the two viable FUSIL bins is shown, along with the likely explanation for these shifts. CL cellular lethal; DL developmental lethal; SV subviable; VP viable with phenotypic abnormalities; VnP viable with no phenotypic abnormalities.

**Supplementary Figure 2 Distributions of tissue specificity values (Tau)**

The distributions of Tau for genes in different FUSIL categories are plotted. From left to right, as indicated, Tau was calculated from either mouse gene expression as measured by ENCODE, mouse gene expression as measured by Cardoso-Moreira et al or human gene expression as measured by Cardoso-Moreira et al. [3]. CL cellular lethal; DL developmental lethal; SV subviable; VP viable with phenotypic abnormalities; VnP viable with no phenotypic abnormalities.

**Supplementary Figure 3 Gene expression patterns**

**(a) Mendelian disease genes across FUSIL bins** Bar plots show the proportion of genes associated with single-gene disorders from PanelApp (green genes, high evidence) for each FUSIL bin. The dashed grey line indicates the proportion of Mendelian disease genes for all protein coding genes according to HGNC [4] **(b) Enrichment analysis of tissues with the highest level of gene expression in Brain, Heart and Cerebellum** Gene expression values across mouse developmental stages were used to compute the tissue with highest levels of gene expression for each gene. Human data, normalised TPM values (max sample (tissue+stage)) from Cardoso-Moreira et al. [3] (see Methods section for computation of OR). The grey dashed line represents OR=1. CL cellular lethal; DL developmental lethal; SV subviable; VP viable with phenotypic abnormalities; VnP viable with no phenotypic abnormalities.

**Supplementary Figure 4 Sequence length and constraint metrics**

**(a) Gene length** Box plots show the distribution of gene lengths across all associated FUSIL genes (Ensembl Genes 113, hsapiens_gene_ensembl dataset). **CDS length** Box plots show the distribution of CDS, the portion of a transcript that excludes UTRs, lengths (longest CDS prioritised per gene) across all associated FUSIL genes (Ensembl Genes 113, hsapiens_gene_ensembl dataset). **Protein length** Box plots show the distribution of protein lengths across all associated FUSIL genes (UniProtKB, Human/9606 data set). Outliers are not shown in the boxplots. **(b) gnomAD LOEUF** Box plots show the distribution of LOEUF scores, obtained from gnomAD v4.0 (https://gnomad.broadinstitute.org/) [5], across all associated FUSIL genes. Genes below the 0.6 threshold can be considered as 'constrained'. **GISMO** Box plots show the distribution of GISMO scores, obtained from [6], across all associated FUSIL genes. **$S_{het}$ (GeneBayes)** Box plots show the distribution of $S_{het}$ scores, obtained from Zeng et al. [7], across all associated FUSIL genes. Higher $S_{het}$ scores indicate more constrained genes. **(c) GISMO-mis** Box plots show the distribution of GIMSO-mis scores, obtained from [6], across all associated FUSIL genes. **gnomAD Z score (missense)** Box plots show the distribution of Z score (missense) scores across all associated FUSIL genes. **AlphaMissense Mean Pathogenicity** Box plots show the distribution of AlphaMissense predicted pathogenicity scores, obtained from Cheng et al. [8], across all associated FUSIL genes. Scores closer to 1 indicate a higher likelihood of a gene being pathogenic. The significance for pairwise comparisons between FUSIL bins across features is shown in **Supplementary Table 3.** CL cellular lethal; DL developmental lethal; SV subviable; VP viable with phenotypic abnormalities; VnP viable with no phenotypic abnormalities.

**Supplementary Figure 5 Diagnostic cases in the 100KGP**

**(a) Distribution of diagnosis across disease categories** Box plots represent the percentage of diagnoses in each FUSIL bin based on the recruited disease category for each patient. Only disease categories ≥2% diagnosis are shown. Data for VnP not shown given the low number of diagnoses in that bin **(b) Phenotypic similarity scores** Pairwise similarity scores between patients based on their associated HPO phenotypes were computed using the PhenoDigm algorithm[9]. The distribution of these scores is plotted for different gene groups, categorised according to FUSIL information and recruited disease category. CL cellular lethal; DL developmental lethal; SV subviable; VP viable with phenotypic abnormalities; VnP viable with no phenotypic abnormalities.

#### Supplementary Figure 1

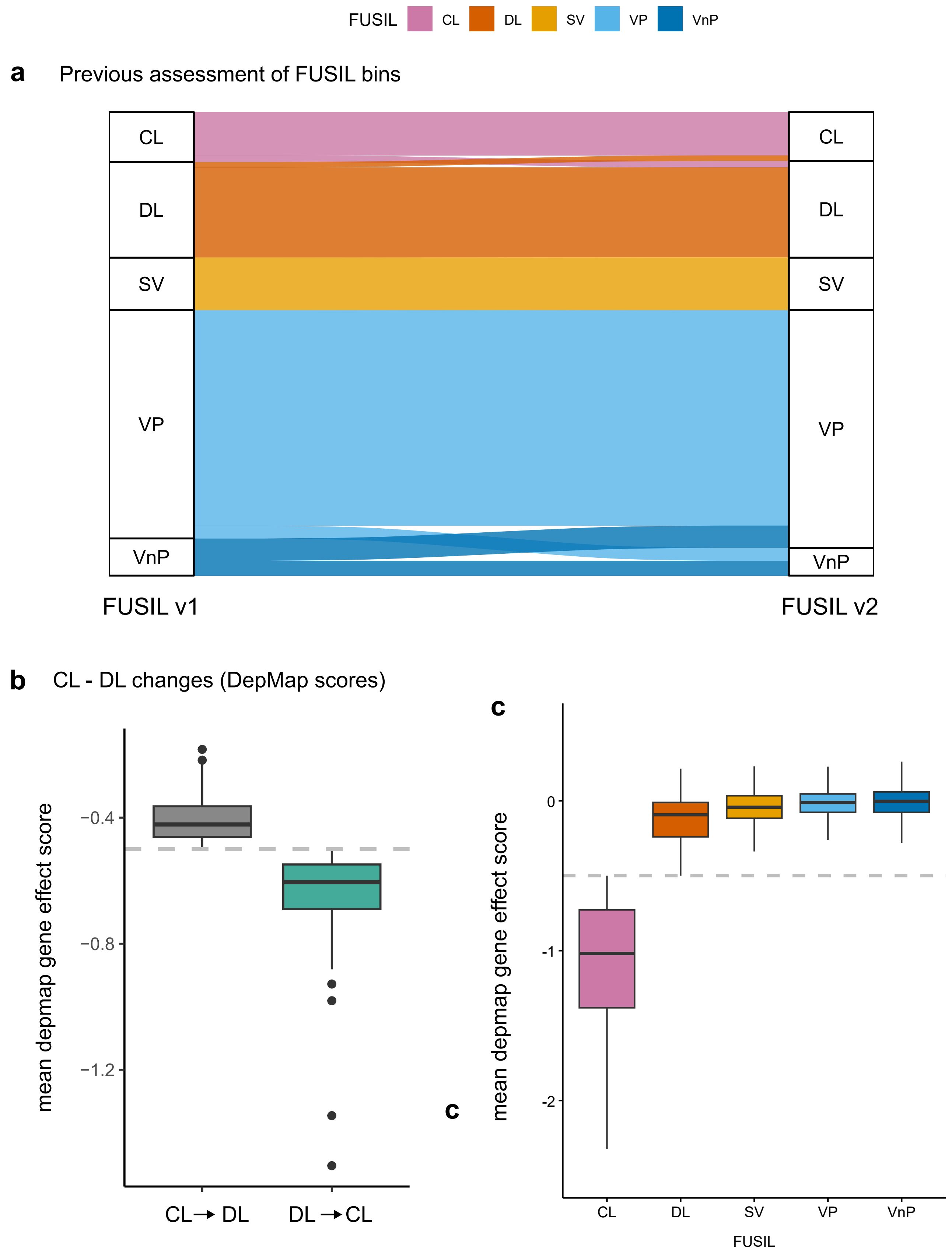


**d** VP - VnP changes

| | | |
|---|---|---|
| VP → VnP | 95 | differences between WT and KO no longer significant (soft- windowing) |
| VnP → VP | 165 | new phenotypes have been assesed |

### Supplementary Figure 2

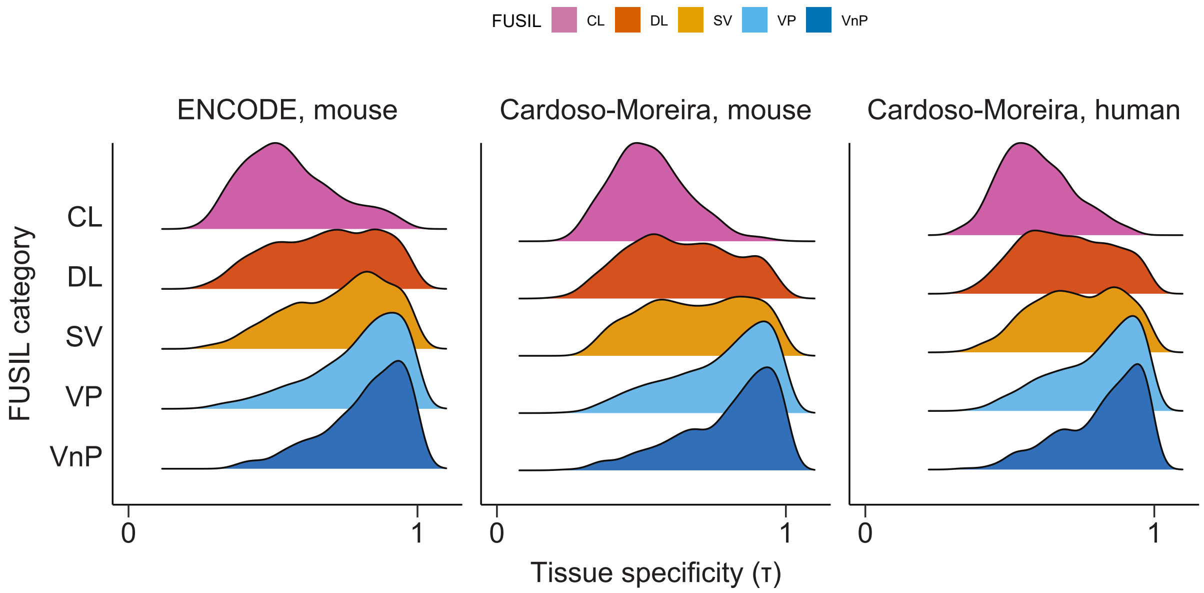

### Supplementary Figure 3

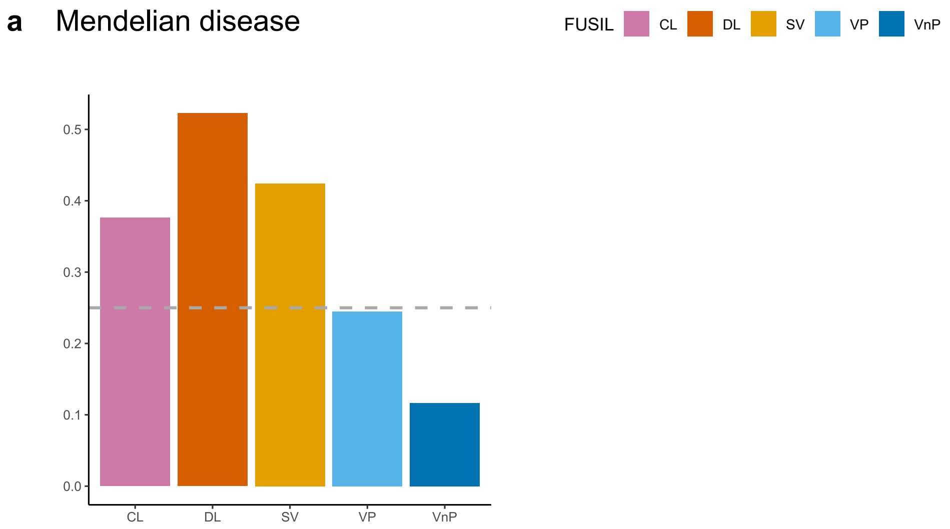


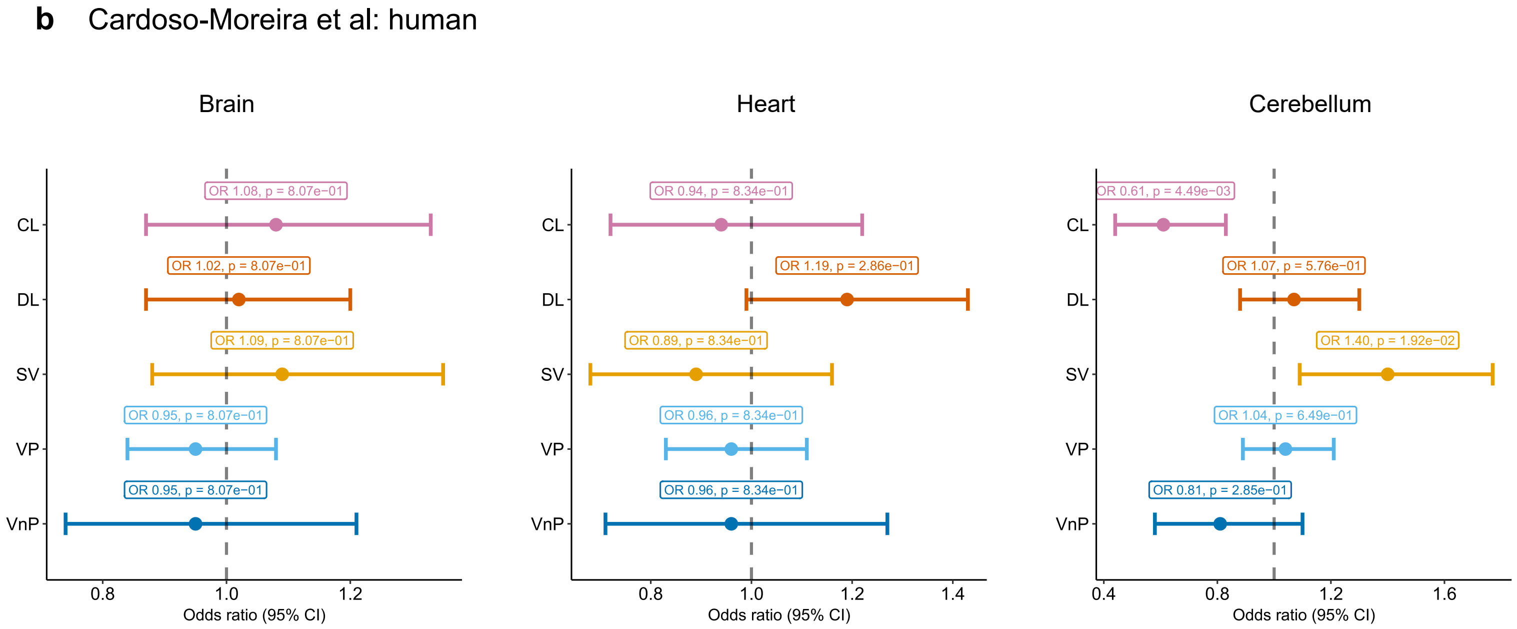

#### Supplementary Figure 4

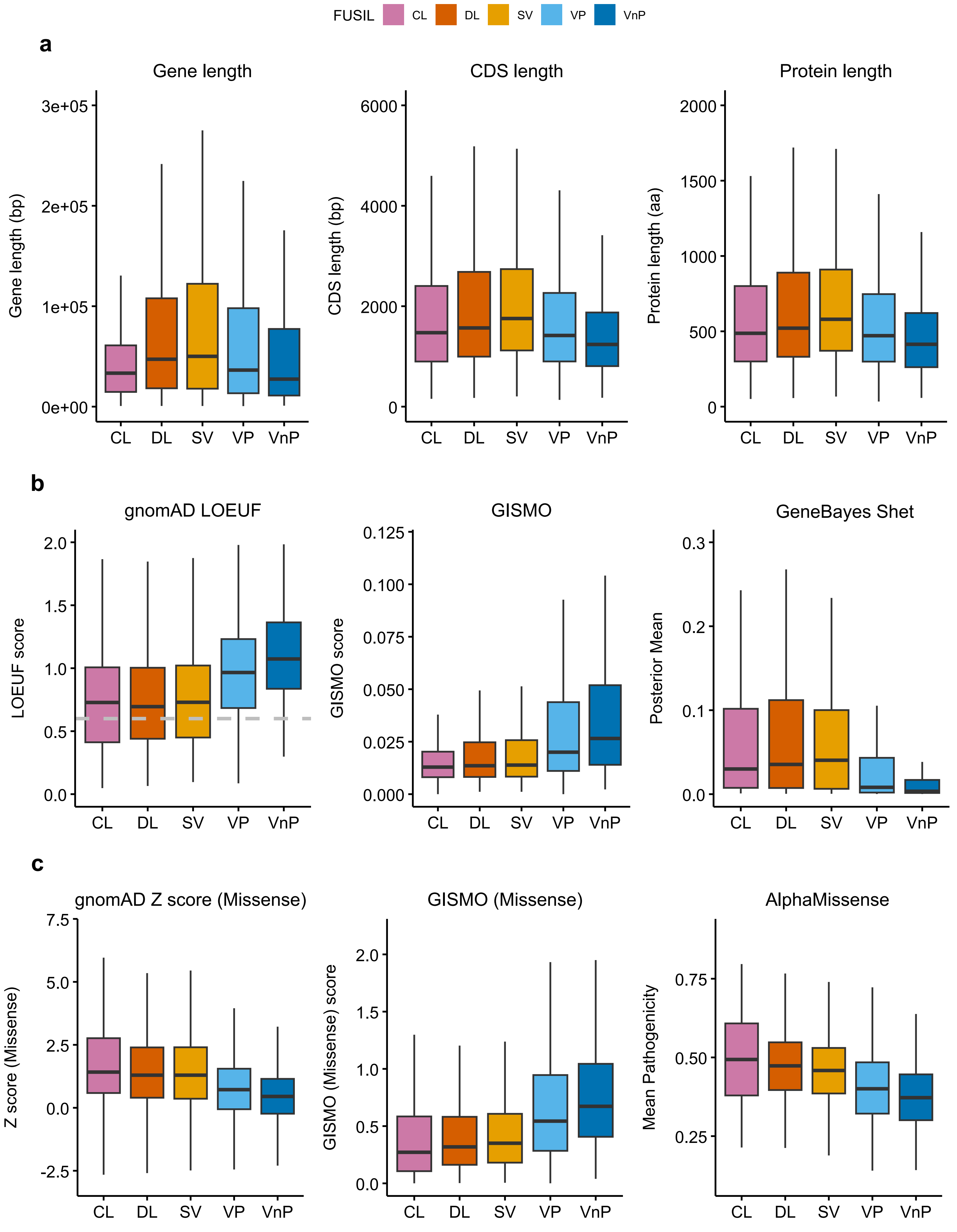

### Supplementary Figure 5

**a** Recruited disease categories

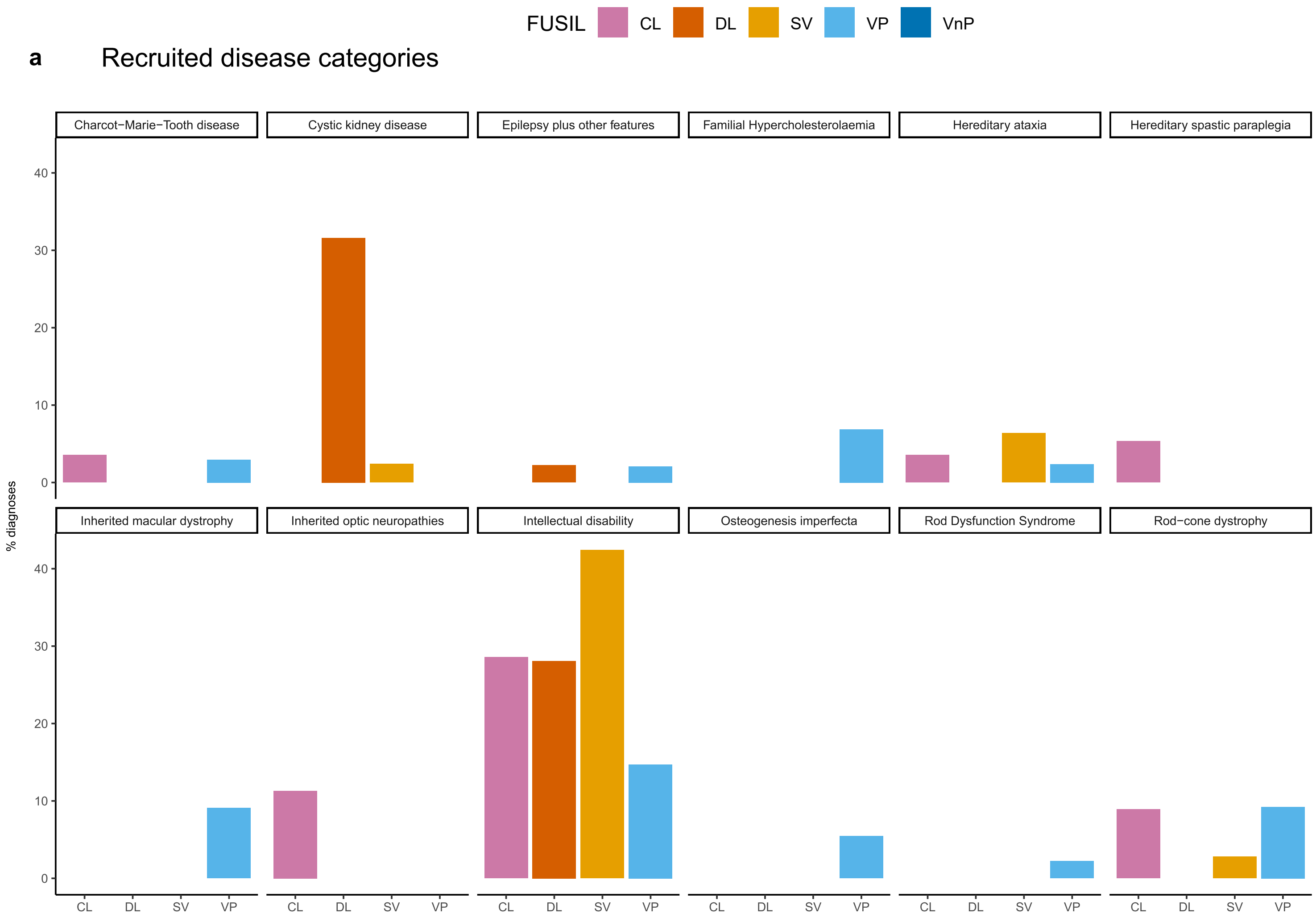


**b** Phenotypic similarity

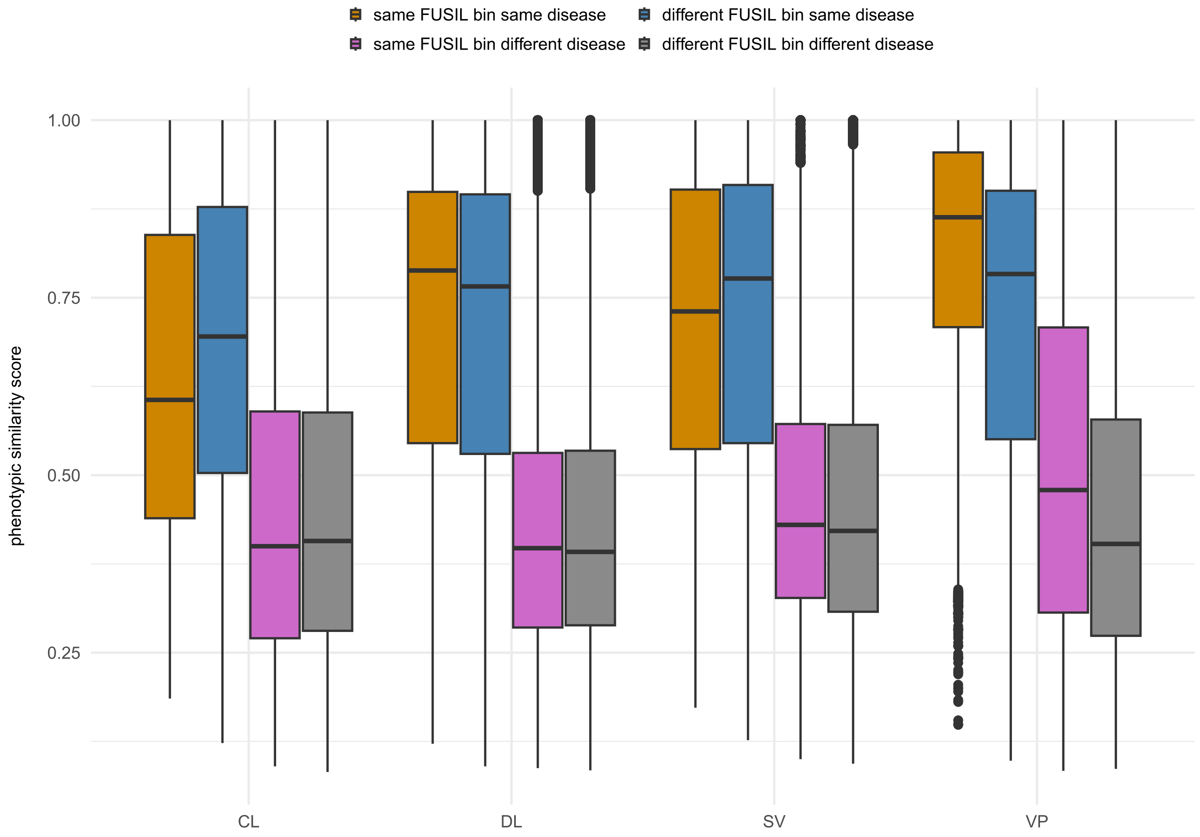

**Supplementary Table 1. Gene length.** BH-adjusted Pvalues (Wilcoxon test, two-sided) for all pairwise comparisons for each feature. See the Methods section for a description of each feature. CL cellular lethal; DL developmental lethal; SV subviable; VP viable with phenotypic abnormalities; VnP viable with no phenotypic abnormalities.

| Comparison | Gene length | Transcript length | CDS length | Protein length |
|---|---|---|---|---|
| **CL vs DL** | 9.5E-11 | 1.9E-08 | 6.7E–03 | 1.1E-2 |
| **CL vs SV** | 9.5E-11 | 2.4E-12 | 1.1E-04 | 1.8E-4 |
| **CL vs VP** | 8.3E-4 | 1.1E-2 | 4.9E-01 | 4.1E-1 |
| **CL vs VnP** | 7.3E-1 | 1.4E-1 | 2.8E-04 | 2.1E-4 |
| **DL vs SV** | 2.4E-1 | 1.1E-2 | 6.1E-02 | 7.4E-2 |
| **DL vs VP** | 1.7E-05 | 1.9E-07 | 8.7E-07 | 7.2E-07 |
| **DL vs VnP** | 8.4E-08 | 7.3E-12 | 3.6E-11 | 3.1E-11 |
| **SV vs VP** | 8.9E-06 | 8.8E-12 | 1.8E-09 | 2.2E-09 |
| **SV vs VnP** | 3.5E-08 | 4.5E-16 | 3.9E-14 | 3.9E-14 |
| **VP vs VnP** | 4.1E-3 | 3.2E-05 | 3.9E-05 | 4E-05 |

**Supplementary Table 2. Sequence conservation metrics.** BH-adjusted Pvalues (Wilcoxon test, two-sided) for all pairwise comparisons for each feature. See the Methods section for a description of each feature. CL cellular lethal; DL developmental lethal; SV subviable; VP viable with phenotypic abnormalities; VnP viable with no phenotypic abnormalities.

| Comparison | phyloP17 way primate | phyloP470 way mammalian | phyloP100 way vertebrate | phastCons17 way primate | phastCons470 way mammalian | phastCons100 way vertebrate |
|---|---|---|---|---|---|---|
| **CL vs DL** | 4.2E-01 | 5.7E-01 | 7.2E-03 | 1.8E-01 | 8.6E-01 | 7.2E-02 |
| **CL vs SV** | 2.1E-01 | 8.6E-02 | 9.4E-05 | 1.9E-01 | 5.4E-01 | 8.1E-03 |
| **CL vs VP** | <2.0E-16 | <2.0E-16 | <2.0E-16 | <2.0E-16 | <2.0E-16 | <2.0E-16 |
| **CL vs VnP** | <2.0E-16 | <2.0E-16 | <2.0E-16 | <2.0E-16 | <2.0E-16 | <2.0E-16 |
| **DL vs SV** | 4.6E-01 | 1.3E-01 | 3.5E-02 | 9E-01 | 5.5E-01 | 1.7E-01 |
| **DL vs VP** | <2.0E-16 | <2.0E-16 | <2.0E-16 | <2.0E-16 | <2.0E-16 | <2.0E-16 |
| **DL vs VnP** | <2.0E-16 | <2.0E-16 | <2.0E-16 | <2.0E-16 | <2.0E-16 | <2.0E-16 |
| **SV vs VP** | <2.0E-16 | <2.0E-16 | <2.0E-16 | <2.0E-16 | <2.0E-16 | <2.0E-16 |
| **SV vs VnP** | <2.0E-16 | <2.0E-16 | <2.0E-16 | <2.0E-16 | <2.0E-16 | <2.0E-16 |
| **VP vs VnP** | 1.6E-07 | 8.2E-09 | 1.2E-07 | 8.7E-08 | 1.6E-07 | 1.1E-06 |

**Supplementary Table 3. Constraint metrics.** BH-adjusted Pvalues (Wilcoxon test, two-sided) for all pairwise comparisons for each feature. See the Methods section for a description of each feature. CL cellular lethal; DL developmental lethal; SV subviable; VP viable with phenotypic abnormalities; VnP viable with no phenotypic abnormalities.

| Comparison | gnomAD LOEUF | gnomAD Z score (Missense) | AlphaMissense | GeneBayes (Shet) | GISMO | GISMO (missense) |
|---|---|---|---|---|---|---|
| **CL vs DL** | 8.0E-01 | 1.6E-02 | 1.8E-03 | 7.2E-01 | 4.2E-02 | 8.1E-04 |
| **CL vs SV** | 3.0E-01 | 2.6E-02 | 9.8E-06 | 4.8E-01 | 1.5E-02 | 1.5E-05 |
| **CL vs VP** | <2.0E-16 | <2.0E-16 | <2.0E-16 | <2.0E-16 | <2.0E-16 | <2.0E-16 |
| **CL vs VnP** | <2.0E-16 | <2.0E-16 | <2.0E-16 | <2.0E-16 | <2.0E-16 | <2.0E-16 |
| **DL vs SV** | 3.0E-01 | 9.5E-01 | 1.4E-02 | 2.1E-01 | 4.1E-01 | 3.5E-02 |
| **DL vs VP** | <2.0E-16 | <2.0E-16 | <2.0E-16 | <2.0E-16 | <2.0E-16 | <2.0E-16 |
| **DL vs VnP** | <2.0E-16 | <2.0E-16 | <2.0E-16 | <2.0E-16 | <2.0E-16 | <2.0E-16 |
| **SV vs VP** | <2.0E-16 | <2.0E-16 | <2.0E-16 | <2.0E-16 | <2.0E-16 | <2.0E-16 |
| **SV vs VnP** | <2.0E-16 | <2.0E-16 | <2.0E-16 | <2.0E-16 | <2.0E-16 | <2.0E-16 |
| **VP vs VnP** | <2.1E-10 | 1.1E-05 | 5.5E-07 | 7E-13 | 3.9E-07 | 3.5E-07 |

**Supplementary Table 4. Candidate variants identified in DL and SV genes in the mouse not linked to Mendelian disorders (*denovo* or top ranked in Exomiser)**

| Disease Category | Gene | FUSIL | Variants (Assembly b38) | Exomiser ranking | Functional class | *De novo* | Additional Disease/ Variants meeting criteria | ClinGen final score # | ClinGen classification | IMPC het knockout mouse phenotypes | Mantis ML-max score | gnomAD v4 LOEUF | Gene family information: PanelApp AUS / GEL | Decipher Research Variants |
|---|---|---|---|---|---|---|---|---|---|---|---|---|---|---|
| Intellectual disability | *HDAC2* | DL | 6:113943407:CCTTT:C *<br>6:113953291:T:G<br>6:113943395:GCTTT:G | 1<br>2<br>1 | frameshift<br>missense<br>frameshift | Y<br>Y<br>Y | Osteogenesis imperfecta (missense) | 11 | **Moderate** | No spontaneous movement, unresponsive to tactile stimuli | 99.5 (DD monoallelic) | 0.37 | *HDAC3, HDAC4, HDAC8* (green, ID panel)<br>*HDAC5* (red, Skeletal dysplasia) AUS | 2 missense (*de novo*) |
| Intellectual disability | *DAPK3* | DL | 19:3964262:C:T<br>19:3964710:T:A | 2<br>3 | missense<br>missense | Y<br>Y | NO | 5 | Limited | hyperactivity | 83.2 (ASD monoallelic) | 1.18 | *DAPK3* (red, Additional findings Paediatric, AUS) | NO |
| Intellectual disability | *LEO1* | DL | 15:51966008:C:T<br>15:51951956:G:T | 11<br>3 | missense<br>missense | Y<br>Y | Classical tuberous sclerosis (missense)<br>Hereditary ataxia (stop gained) | 5.5 | Limited | NO | 93.5 (ASD monoallelic) | 0.488 | ***LEO1*** (amber, ID, AUS) | 2 frameshift, 2 missense (*de novo*) |
| Epilepsy (plus other features) | *LHX6* | SV | 9:122213632:A:G *<br>9:122228305:A:G<br>9:122226503:G:T | 2<br>1<br>10 | start lost<br>splice donor variant<br>splice region variant | | Intellectual disability (splice region variant)<br>Intracerebral calcification disorders (start lost) | 9 | **Moderate** | decreased locomotor activity | 96.7 (DD monoallelic) | 0.594 | *LHX2* (green, Autism), *LHX3, LHX4* (green, Growth failure)<br>*LHX8* (green, Primary ovarian insufficiency) AUS | NO |
| Dilated cardiomyopathy | *GJC1* | DL | 17:44805723:C:T | 2 | missense | | Congenital neutropaenia (missense)<br>Early onset dystonia (missense) | 5.1 | Limited | Abnormal pericardium morphology (hom, E9.5) | - | 0.348 | *GJC2* (green in Fetal anomalies and other panels) AUS | NO |

Rankings Exomiser: (*HDAC2:* ***1*,*2*,*1***,*2); (DAPK3:* ***2*,*3***); (*LEO1:* ***11*,*3***,*6*,*1); (LHX6:* **2**,**1**,**10**,8,9); (*GJC1*: **2**,4,9). CL: cellular lethal; DL: developmental lethal; IMPC: International Mouse Phenotyping Consortium; E: embryonic day; het: heterozygous; hom: homozygous; AUS: PanelAPP Australia; GEL: Genomics England PanelApp; ID: Intellectual disability; ASD: Autism spectrum disorder; DD: Developmental disorder. * Exomiser result count > 1.

### Breakdown of ClinGen score:

| Gene | ClinGen genetic score (0.1 or 0.5 missense, 1.5 or 2 LoF) | ClinGen gene function score | ClinGen expression score | ClinGen PPI score | ClinGen animal model score | ClinGen final score | ClinGen classification |
|---|---|---|---|---|---|---|---|
| *HDAC2* | 5 | 2 | 1 | 1 | 2 | 11 | **Moderate** |
| *DAPK3* | 1 | 1 | 1 | 1 | 1 | 5 | Limited |
| *LEO1* | 1.5 | 2 | 1 | 1 | 0 | 5.5 | Limited |
| *LHX6* | 4 | 1 | 1 | 1 | 2 | 9 | **Moderate** |
| *GJC1* | 0.1 | 2 | 1 | 0 | 2 | 5.1 | Limited |

**Supplementary Table 5. ENCODE files information.** See the Methods section.

| Files | Data access |
|---|---|
| ENCFF554PCI, ENCFF876OPZ, ENCFF322QGD, ENCFF785MZJ, ENCFF557PQT, ENCFF184FWR, ENCFF722NZT, ENCFF270DCV, ENCFF484FBW, ENCFF680TRV, ENCFF304ILZ, ENCFF343KIR, ENCFF386ZTW, ENCFF532MRQ, ENCFF867NEL, ENCFF449ACY, ENCFF836WUM, ENCFF932AJG, ENCFF578AKP, ENCFF867IVC, ENCFF457PAJ, ENCFF794PWS, ENCFF954BEO, ENCFF042VCB, ENCFF361QMV, ENCFF606UHO, ENCFF787VRY, ENCFF384FTH, ENCFF851KEG, ENCFF952IQJ, ENCFF899QOE, ENCFF238YUA, ENCFF330SEA, ENCFF670AQP, ENCFF319UHW, ENCFF148BEQ, ENCFF243EAY, ENCFF805SYI, ENCFF734XAZ, ENCFF785KAJ, ENCFF484QWQ, ENCFF521YOL, ENCFF720JIN, ENCFF145PTV, ENCFF151ERW, ENCFF240PJE, ENCFF649WEQ, ENCFF238RTX, ENCFF081SJL, ENCFF865VYW, ENCFF173NFQ, ENCFF550DPX, ENCFF960KJV, ENCFF620ZGA, ENCFF264KWG, ENCFF979TLF, ENCFF406BLP, ENCFF997ZPU, ENCFF609AAY, ENCFF868TGE, ENCFF633HRL, ENCFF770SOB, ENCFF159DWP, ENCFF889VIN, ENCFF908DRV, ENCFF745ACD, ENCFF540ERL, ENCFF858ZON, ENCFF343KWN, ENCFF029UVS, ENCFF080PBH, ENCFF611VKF, ENCFF804NPY, ENCFF971KKK, ENCFF126VCW, ENCFF915GEL, ENCFF928MQD, ENCFF727RIO, ENCFF698VUU, ENCFF110ZFH | ENCODE (Mouse) [https://www.encodeproject.org/]<br><br>Data accessed 17.01.25<br><br>ENCODE Project Consortium *et al.* Expanded encyclopaedias of DNA elements in the human and mouse genomes. *Nature* **583**, 699–710 (2020). He, P. *et al.* The changing mouse embryo transcriptome at whole tissue and single-cell resolution. *Nature* **583**, 760–767 (2020). |
